# Advancing precision care in pregnancy through an actionable fetal findings list

**DOI:** 10.1101/2024.09.26.24314442

**Authors:** Jennifer L. Cohen, Michael Duyzend, Sophia M. Adelson, Julie Yeo, Mark Fleming, Rebecca Ganetzky, Rebecca Hale, Deborah M. Mitchell, Sarah U. Morton, Rebecca Reimers, Amy Roberts, Alanna Strong, Weizhen Tan, Jay R. Thiagarajah, Melissa A. Walker, Robert C. Green, Nina B. Gold

## Abstract

The use of genomic sequencing (GS) for prenatal diagnosis of fetuses with sonographic abnormalities has grown tremendously over the past decade. Fetal GS also offers an opportunity to identify incidental genomic variants that are unrelated to the fetal phenotype, but may be relevant to fetal and newborn health. There are currently no guidelines for reporting incidental findings from fetal GS.

In the United States, GS for adults and children is recommended to include a list of “secondary findings” genes (ACMG SF v3.2) that are associated with disorders for which surveillance or treatment can reduce morbidity and mortality. The genes on ACMG SF v3.2 predominantly cause adult-onset disorders. Importantly, many genetic disorders with fetal and infantile onset are actionable as well.

A proposed solution is to create a “fetal actionable findings list,” which can be offered to pregnant patients undergoing fetal GS or eventually, as a standalone cell-free fetal DNA screening test. In this integrative review, we propose criteria for an actionable fetal findings list, then identify genetic disorders with clinically available or emerging fetal therapies, and those for which clinical detection in the first week of life might lead to improved outcomes. Finally, we synthesize the potential benefits, limitations, and risks of an actionable fetal findings list.

## Introduction

The clinical use of genomic sequencing (GS) for prenatal diagnosis of fetuses with sonographic abnormalities has grown tremendously in recent years. The International Society of Prenatal Diagnosis (ISPD) recommends offering fetal GS to patients with pregnancies affected by a major single anomaly, multiple anomalies, or with a history of an undiagnosed fetus or child with a congenital anomaly,^1^ likely affecting up to 2-3% of pregnancies.^2^ The diagnostic yield of GS varies by indication, ranging from 2% for isolated increased nuchal translucency to 53% for skeletal abnormalities.^3^

In the United States, indication-based genome sequencing for children and adults includes the optional analysis of a list of “secondary findings” genes recommended by the American College of Medical Genetics (ACMG) (ACMG SF v3.2).^4^ These genes are predominantly associated with adult-onset cardiac, cancer, and inherited metabolic disorders (IMD). Secondary findings have been identified in at least 1-3% of adults.^5–8^ Once detected, these disorders can often be managed with medication, dietary changes, or long-term surveillance aimed at improving morbidity and mortality in affected individuals.^9^ The ACMG secondary findings list is recommended both for adults and children undergoing GS, however, professional organizations differ in their recommendations on reporting secondary findings for adult-onset conditions in children. While the recommendations of organizations such as Genomics England^10^ generally align with ACMG, the European Society of Human Genetics and others argue that it is premature to screen for later-onset conditions in children.^11^

The ACMG SF v3.2 recommendations do not apply to fetuses, and guidance regarding the reporting of incidental findings from fetal GS remains unclear.^12^ An ACMG Points to Consider document states that, “prenatal exome sequencing analysis could be limited to the reporting of variants in genes associated with the ultrasound findings,” while also recommending that “highly penetrant pathogenic variants detected in genes unrelated to the fetal phenotype, but known to cause moderate to severe childhood onset disorders, are recommended to be reported.”^13^ The ISPD suggests that secondary findings analyzed in fetal GS might include “moderate to severe childhood conditions,” but does not provide specific guidance on which genes to include.^1^ Reporting practices vary across clinical labs, and pregnant patients are not routinely offered a choice regarding the type of genomic findings they receive.

Of note, many pregnancies affected by actionable monogenic conditions show no sonographic abnormalities, or the abnormalities are too subtle to detect with current imaging technology.^14,15^ Recent studies have demonstrated that 0.6-2.7% of sonographically normal fetuses harbor pathogenic or likely pathogenic variants (PLPV) expected to cause genetic disease^16–19^ and 1.85% to 9.4% of infants have PLPV associated with a monogenic childhood-onset disorder,^20,21^ including IMD, cardiomyopathies, and syndromic intellectual disability disorders.

Over 700 genetic disorders are now treatable with dietary changes, medication, hematopoietic stem cell transplantation, solid organ transplantation, or gene therapies.^22^ The number of *in utero* therapies for genetic disorders is rapidly expanding,^23^ supported by clinical trials, case reports, and animal model studies. Many of these disorders do not present with ultrasound findings, meaning that the benefits of prenatal therapies can only be realized by patients with known family history or carrier status.^24^ However, most countries lack a uniform approach to carrier screening^25^ and the genes included in these panels vary widely, with none addressing *de novo* disorders in the fetus.^26,27^

Shortly after birth, newborn screening (NBS) identifies many severe, treatable genetic disorders in infants. However, many of the disorders included in NBS programs can cause morbidity or mortality shortly after birth, before the receipt of results at 5 to 7 days of life.^28^ Prenatal diagnosis of these disorders may allow for improved care during the perinatal period, including: appropriate labor and delivery planning, mobilization of relevant medical teams, and the acquisition of specialized medical formulas, medications, or other therapeutics needed for immediate intervention.

A prior commentary by Gold, *et al* suggested offering pregnant patients who are undergoing fetal GS the optional analysis of an “actionable fetal findings list.”^29^ Now that fetal exome sequencing using cell-free fetal DNA (cfDNA) is feasible,^30–32^ this list could also serve as the foundation for a non-invasive cfDNA test available to all pregnant patients. This list is not intended to replace NBS or diagnostic GS for infants, but would enhance reproductive options and management capabilities of conditions not typically identified during pregnancy or the immediate perinatal period.

We used an integrative review approach to propose criteria for an actionable fetal findings list, then identified genetic disorders with clinically available or emerging fetal therapies, and those for which clinical detection in the first week of life might lead to improved outcomes. Finally, we synthesized the potential benefits, limitations, and risks of an actionable fetal findings list.

## Methods

### Study design

The integrative review includes five stages: problem identification, literature search, data evaluation, data analysis, and presentation.^33,34^ This approach allows for the use of several study designs, with the aim of generating new frameworks or ideas.^33,34^

### Selection criteria for genes associated with actionable fetal disorders

We established selection criteria (Table 1) and created three lists: genetic disorders with available or experimental *in utero* therapies (Table 2) and genetic disorders for which prenatal diagnosis could plausibly improve outcomes in the first week of life (Table 3). Genetic disorders with *in utero* therapies that have been used in animal models have been listed separately, as these may be future candidates for an actionable fetal findings list (Table S1).

**Table 1.**
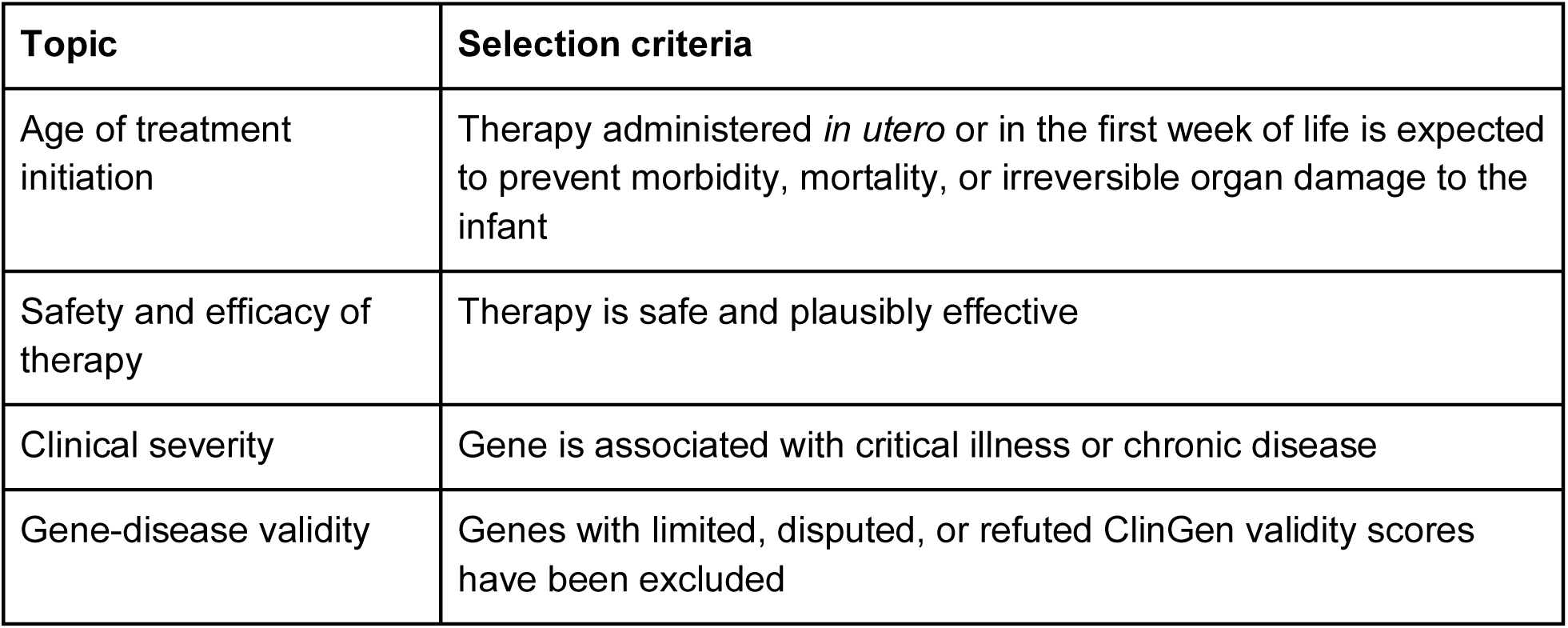
Selection criteria for genes associated with actionable fetal disorders.

**Table 2.**
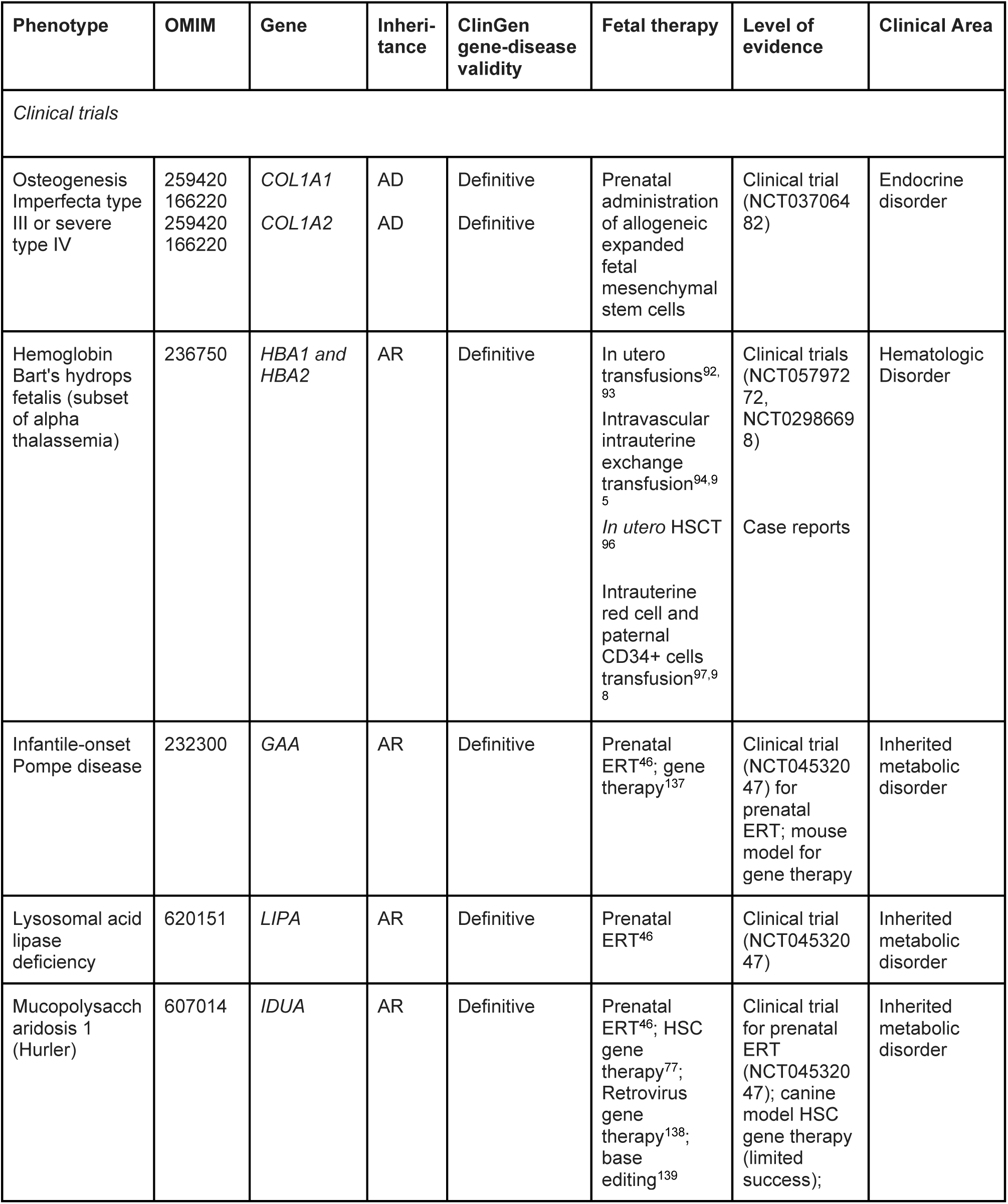
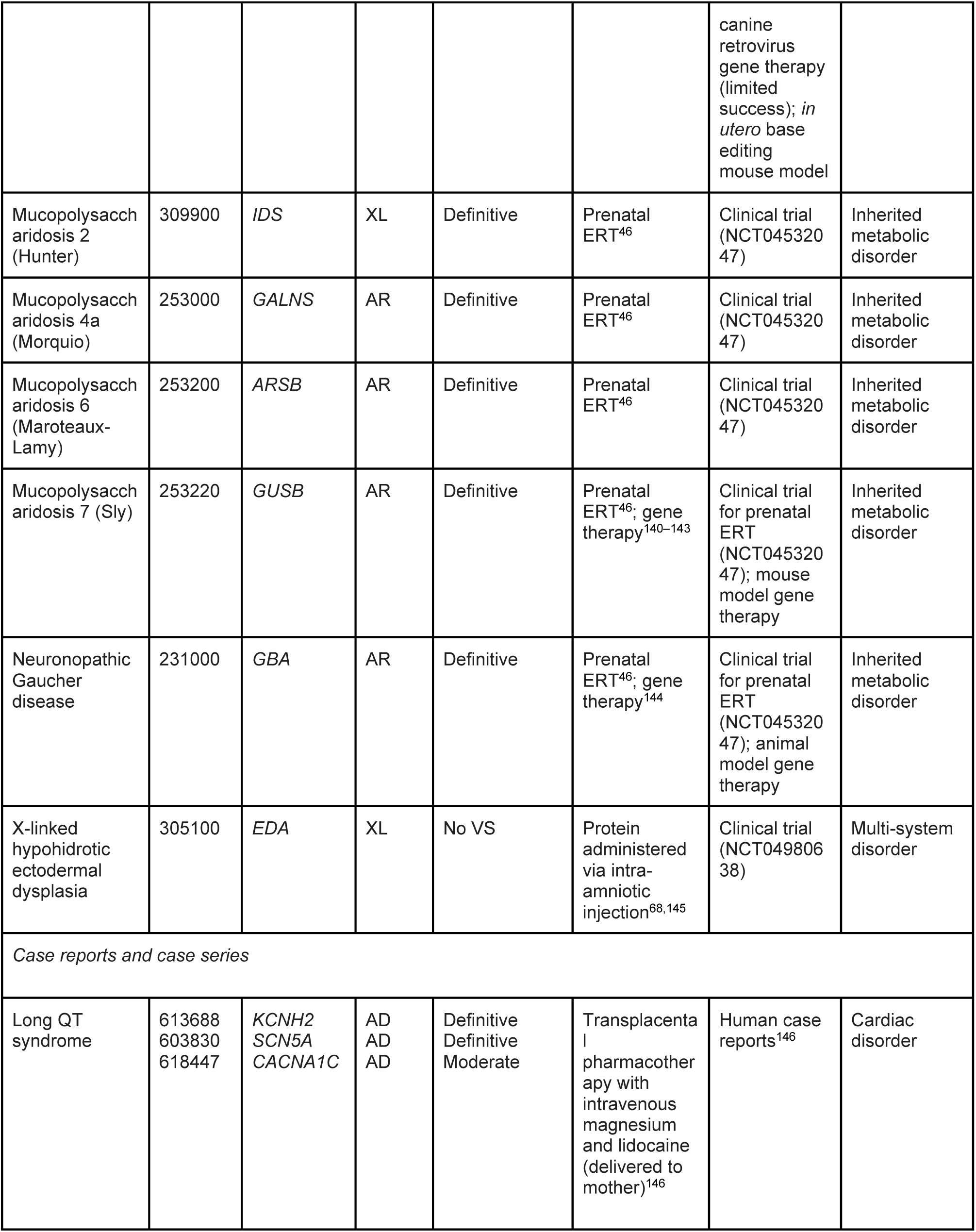
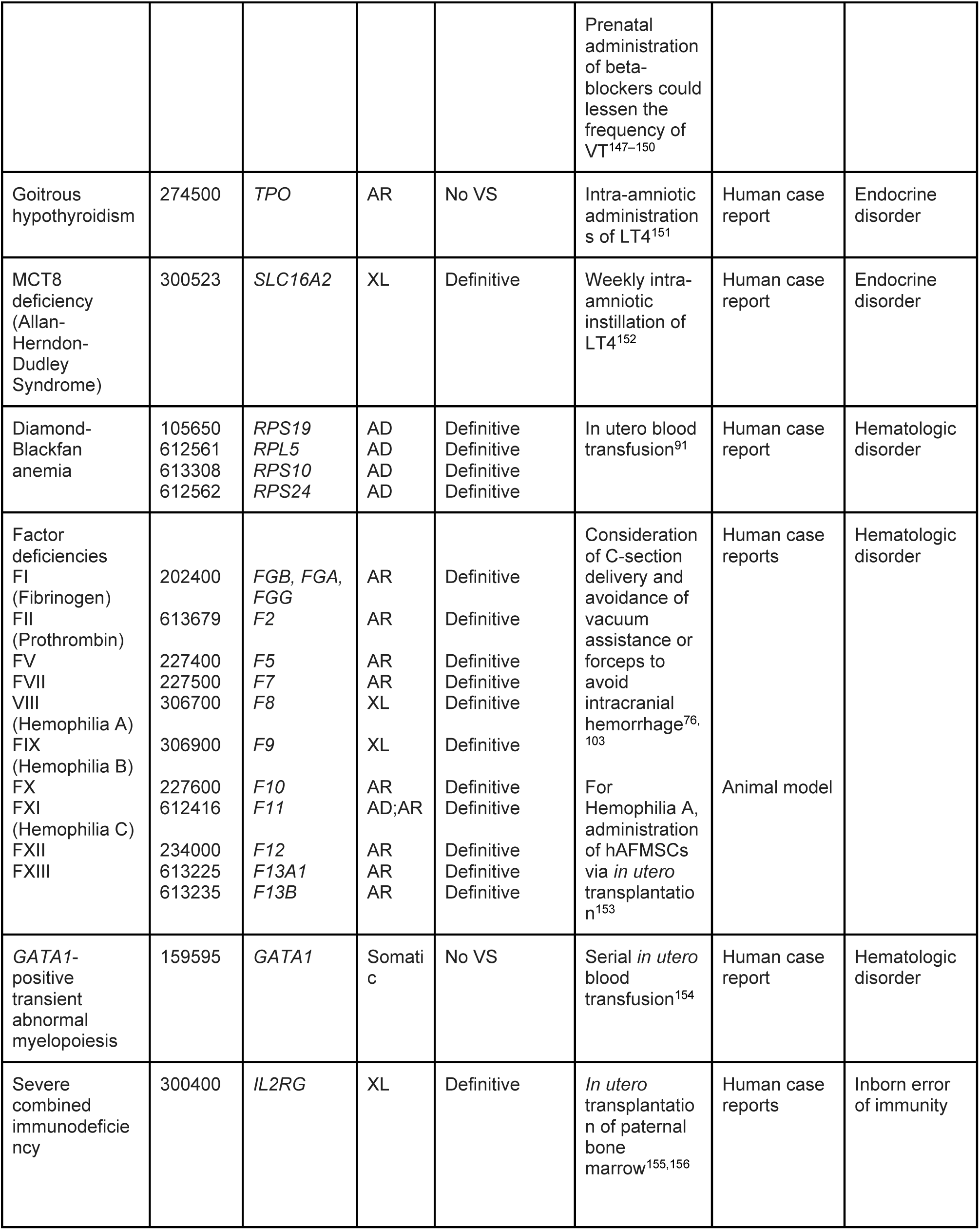
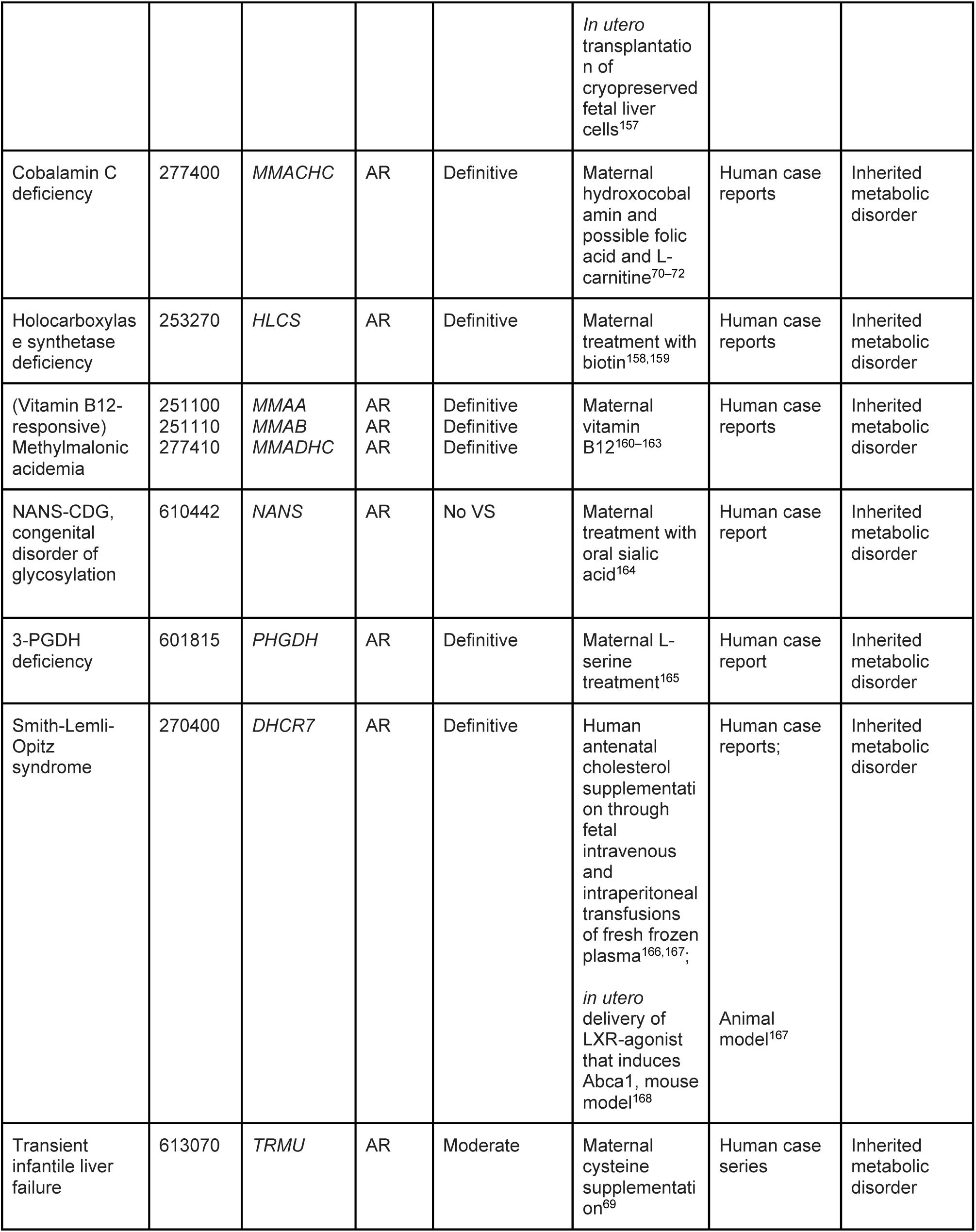
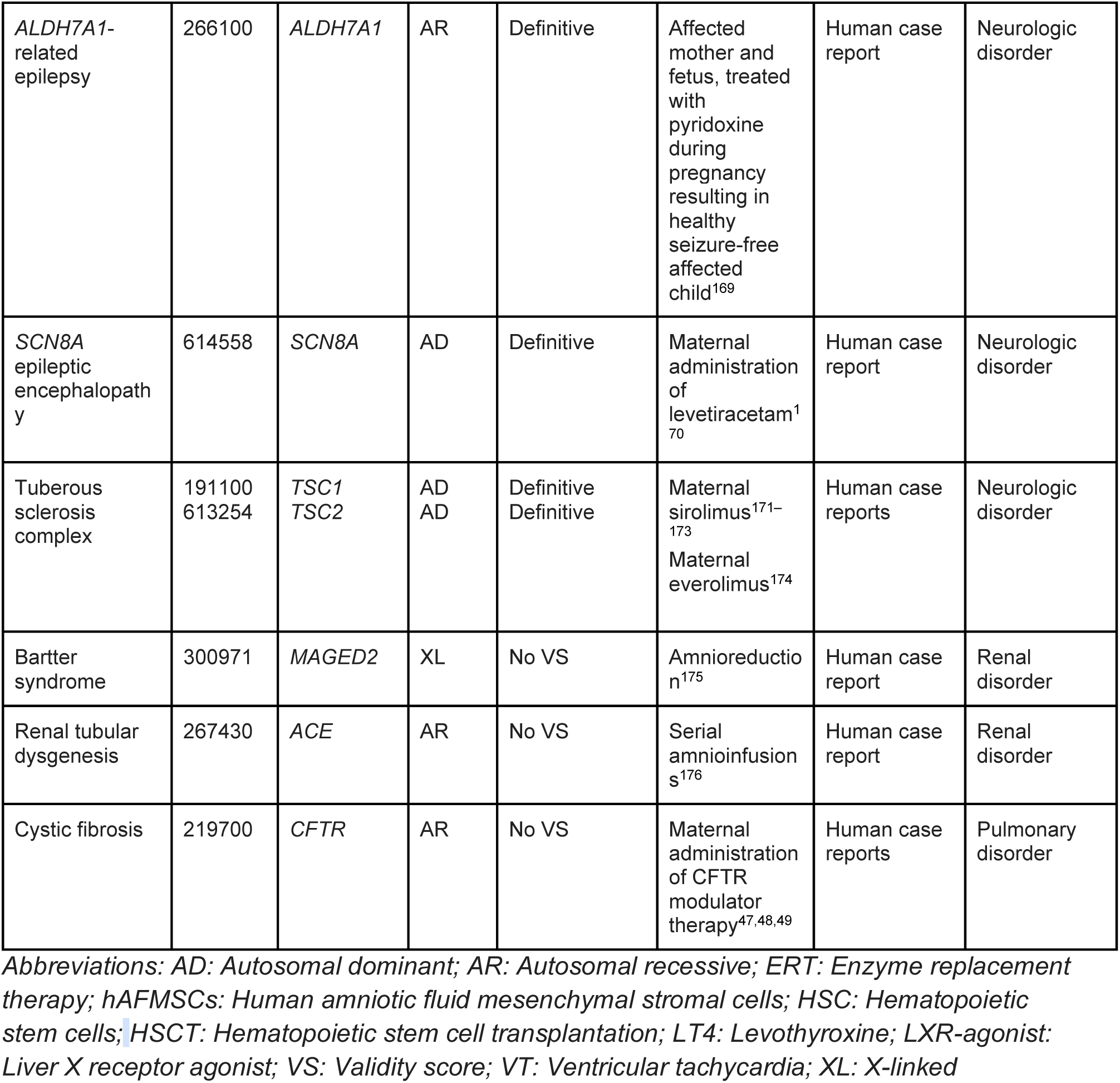
Genes associated with disorders with *in utero* fetal therapies in clinical trials and case reports (n = 53).

**Table 3.**
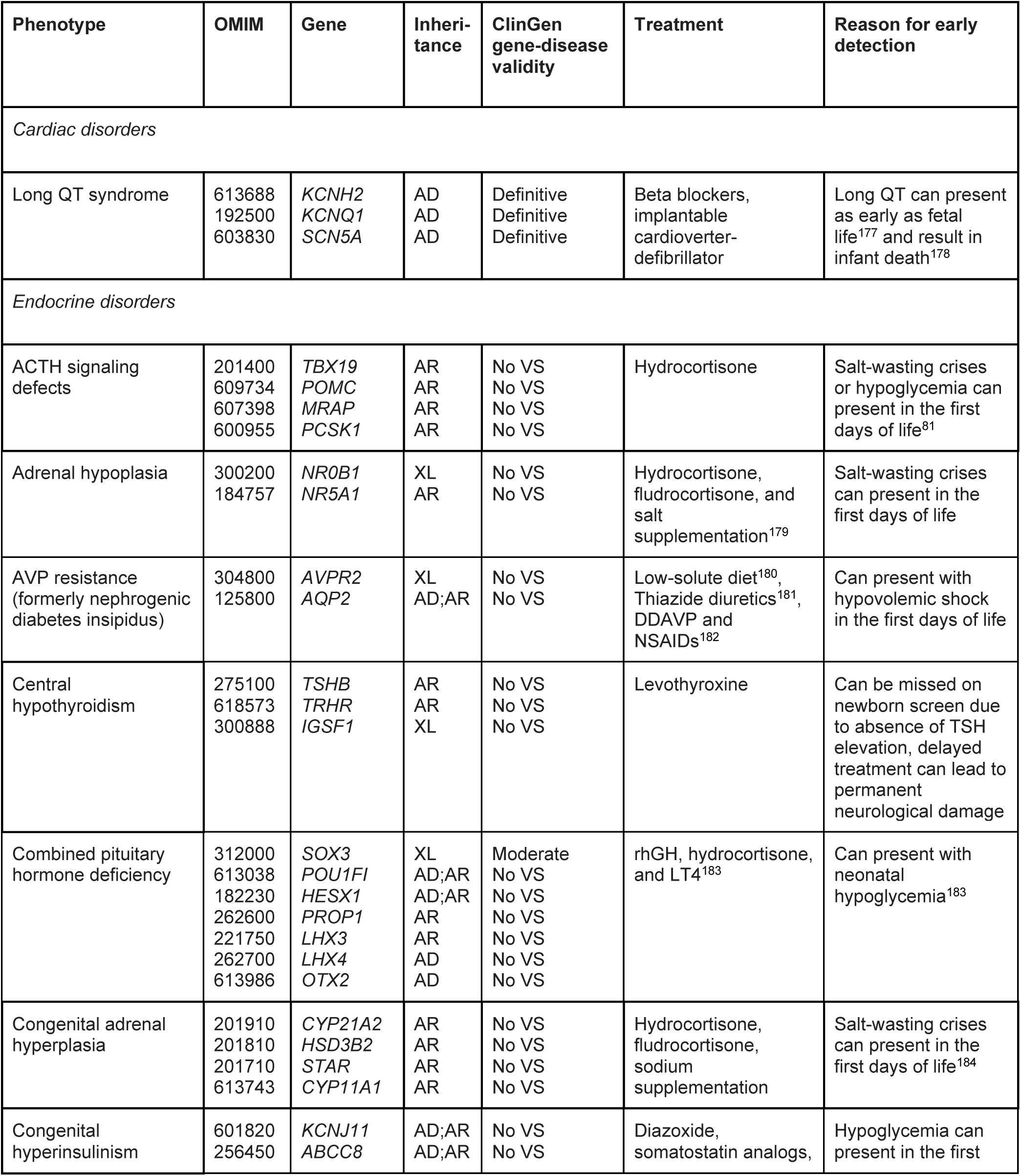
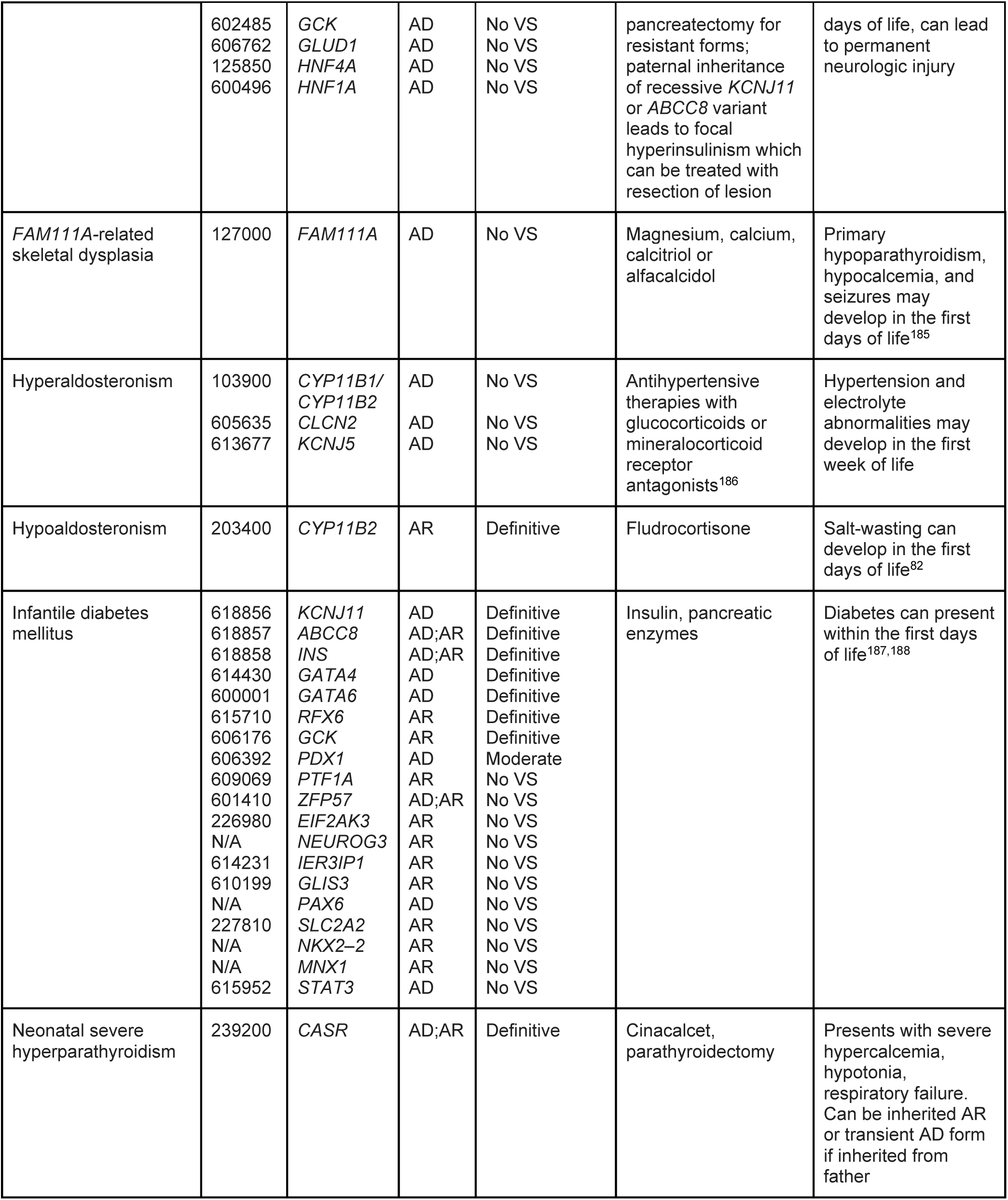
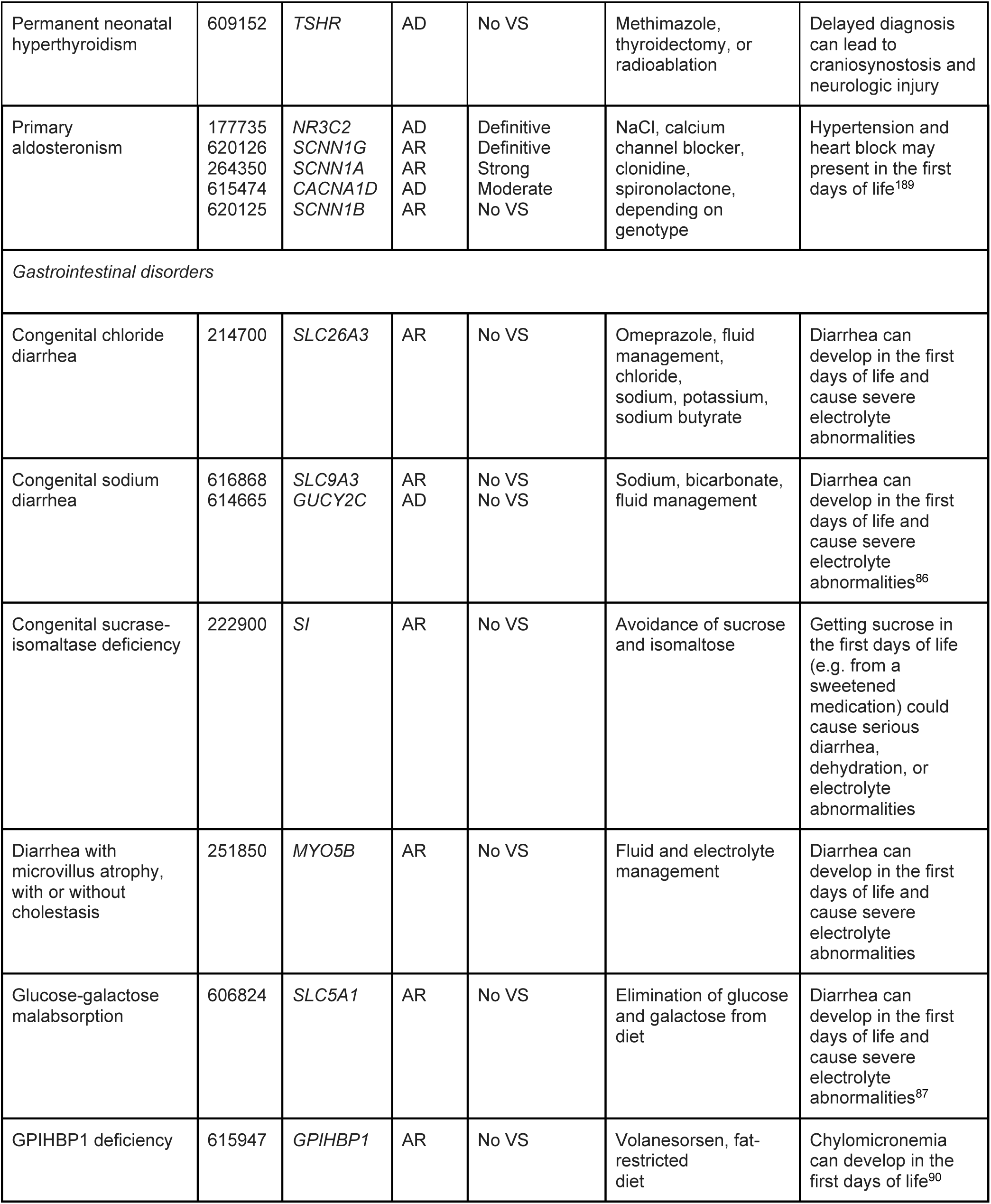
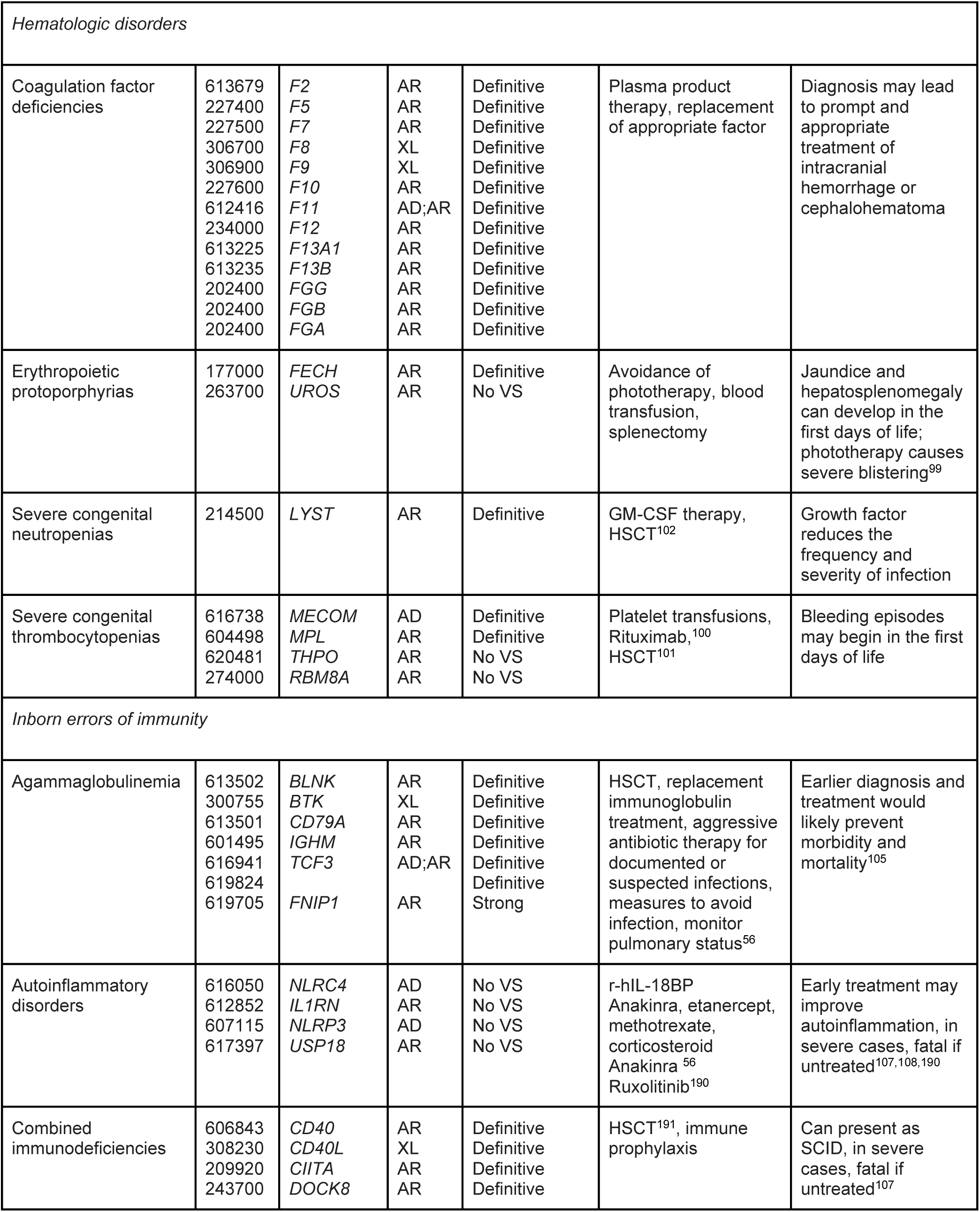
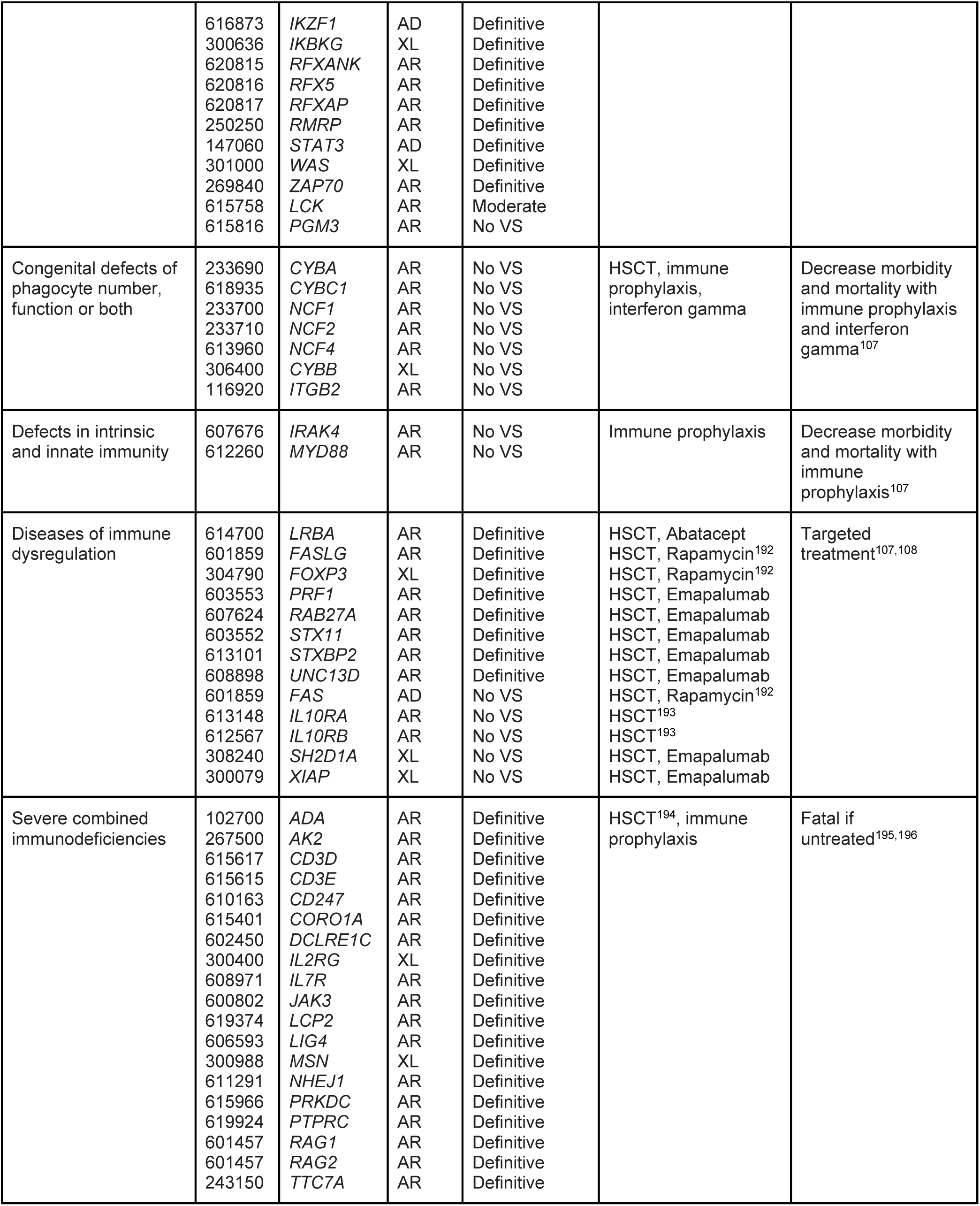
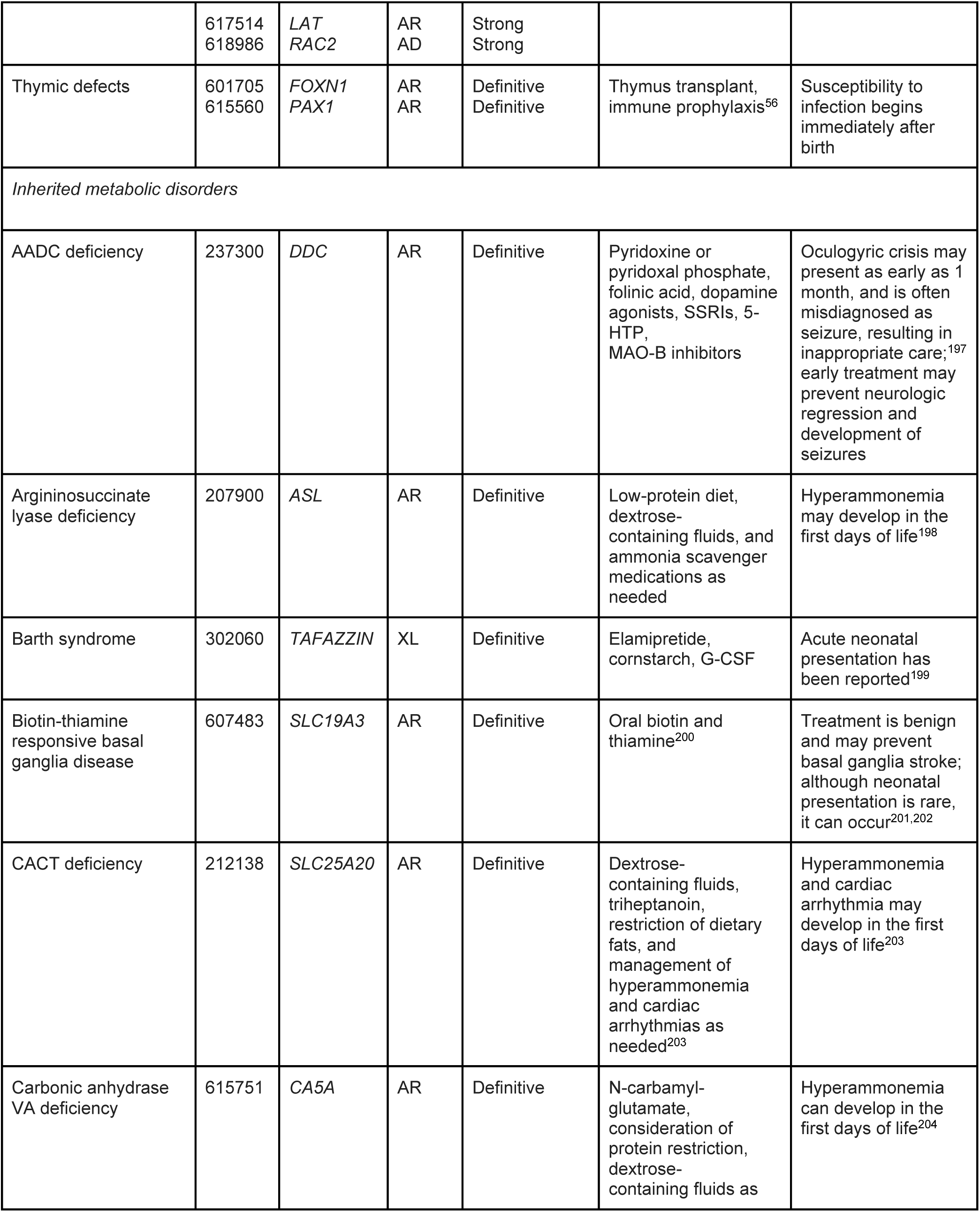
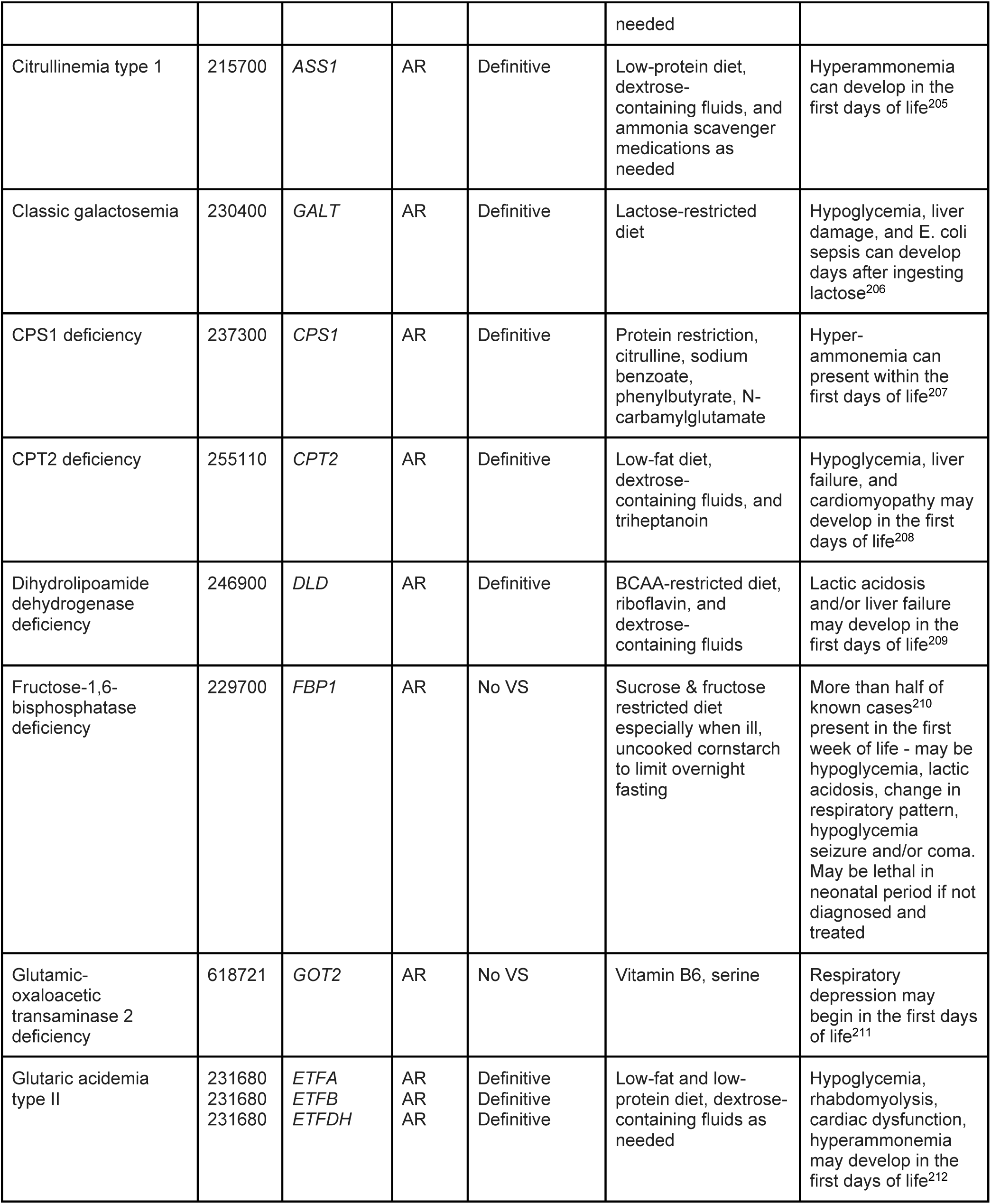
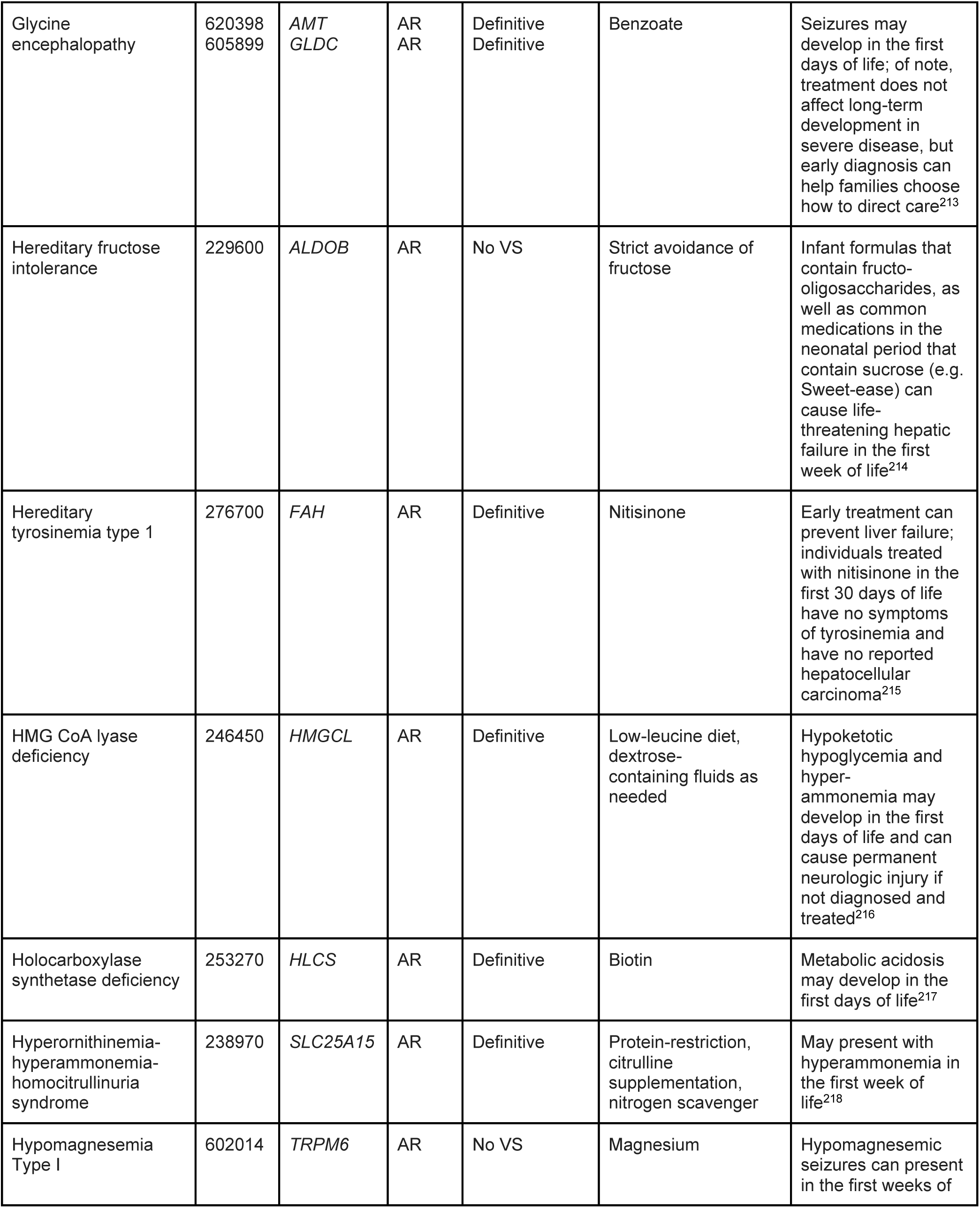
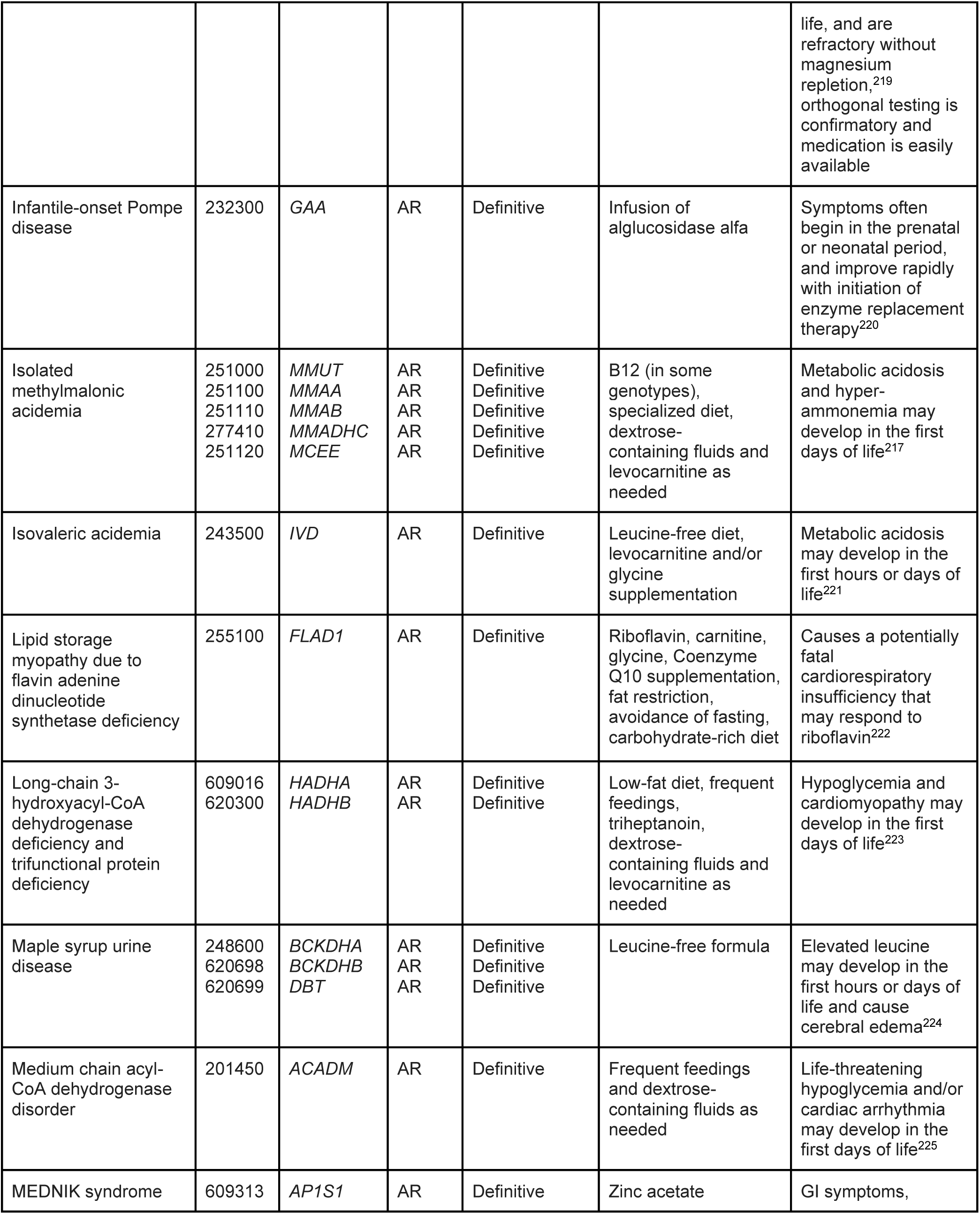
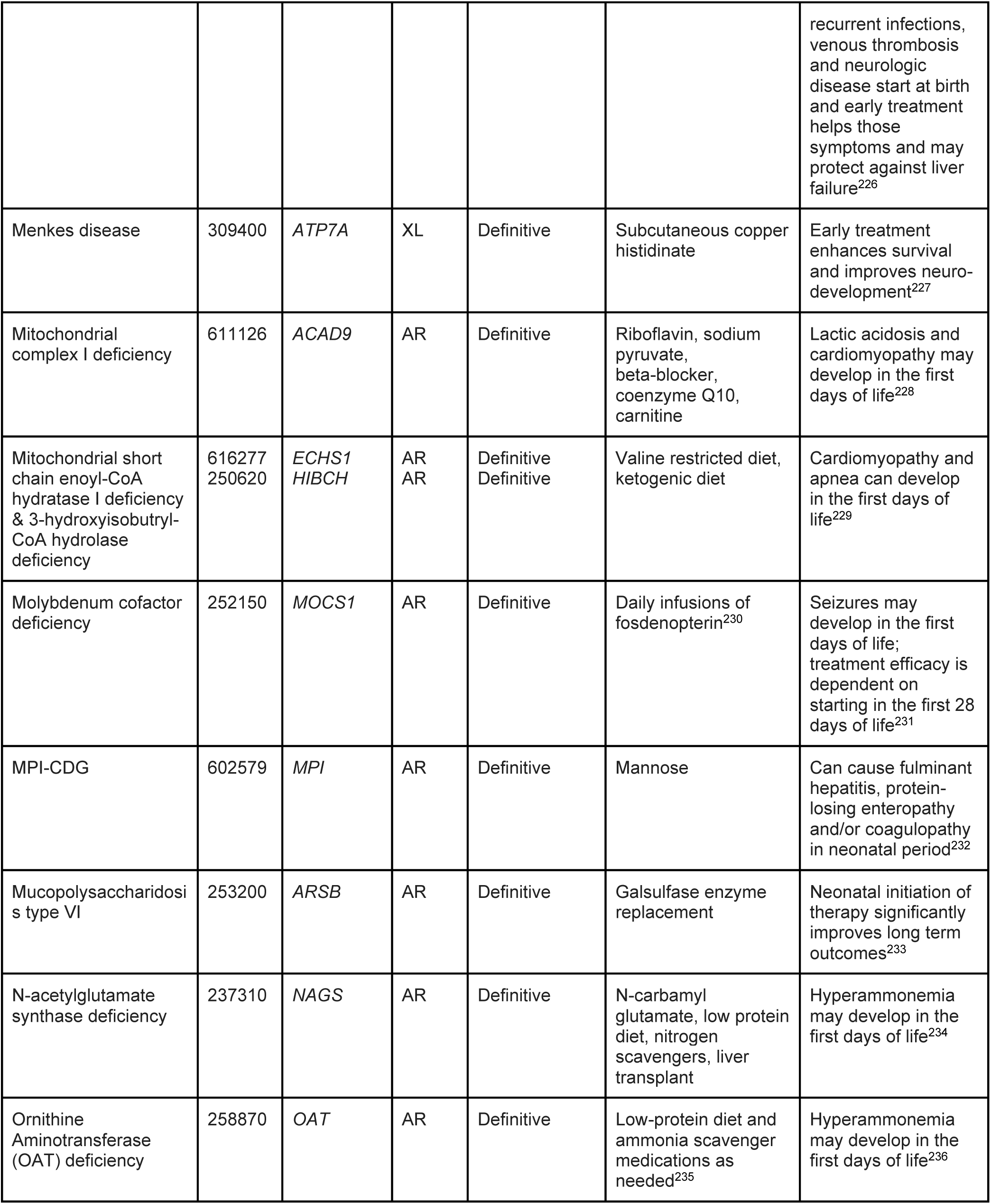
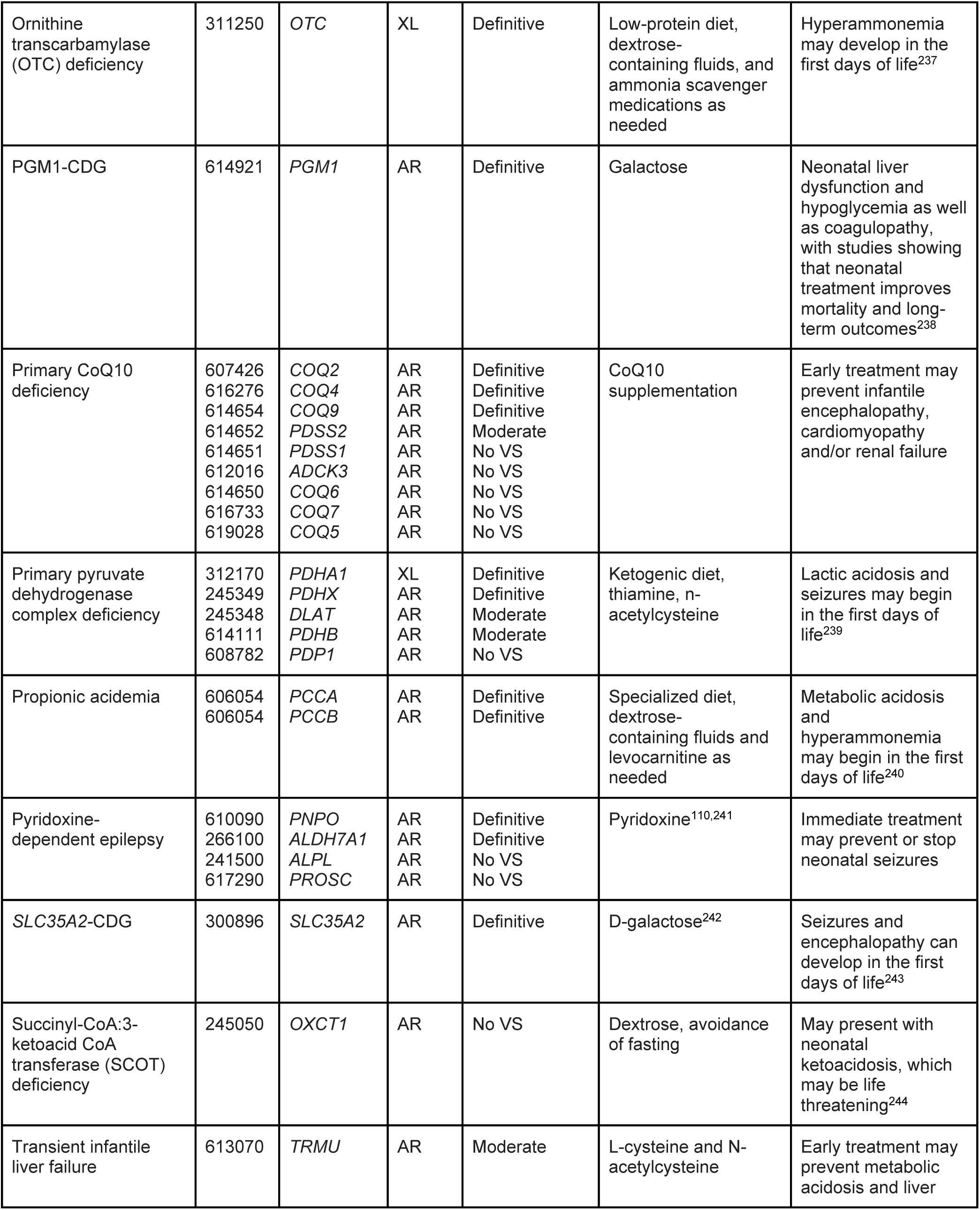
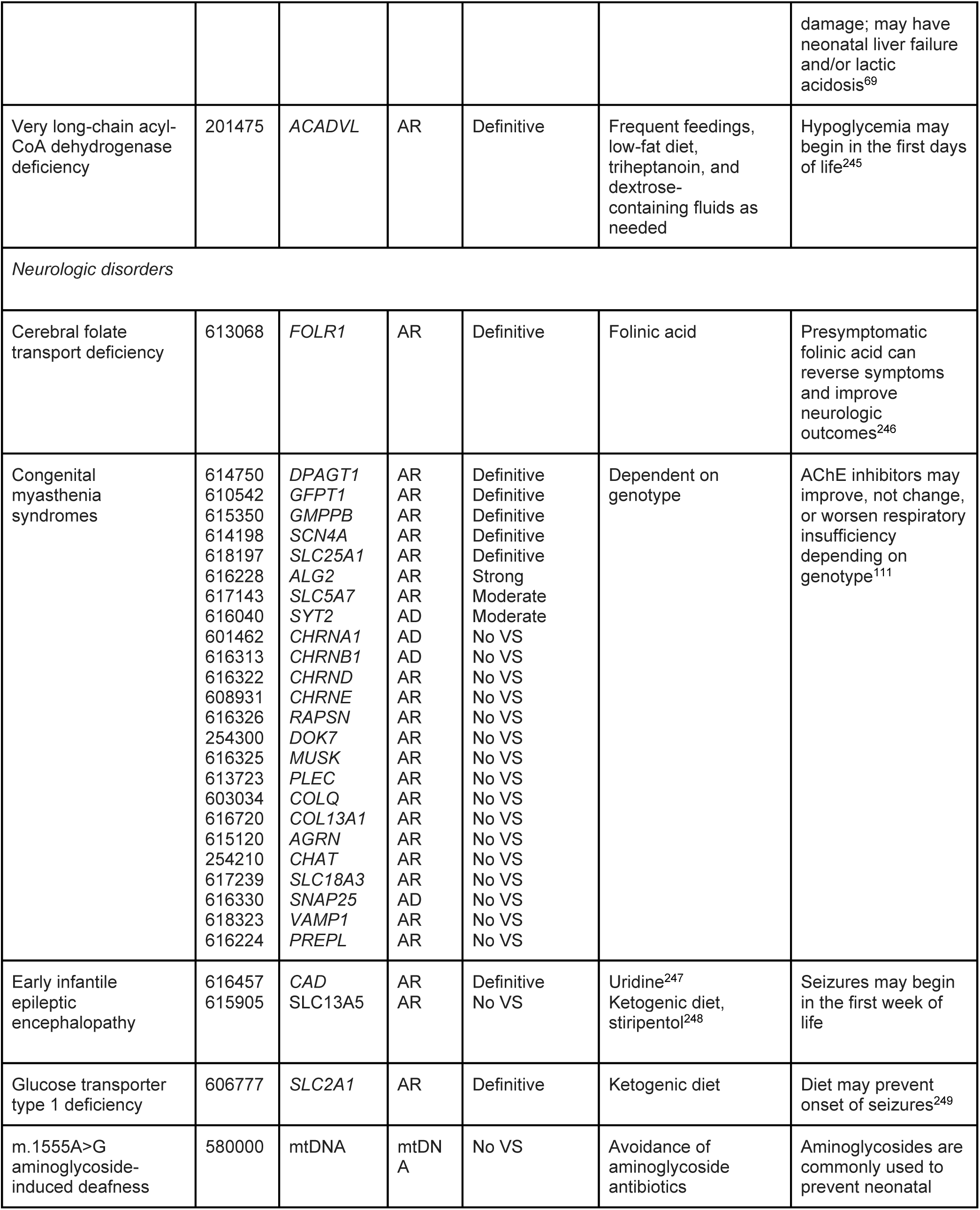
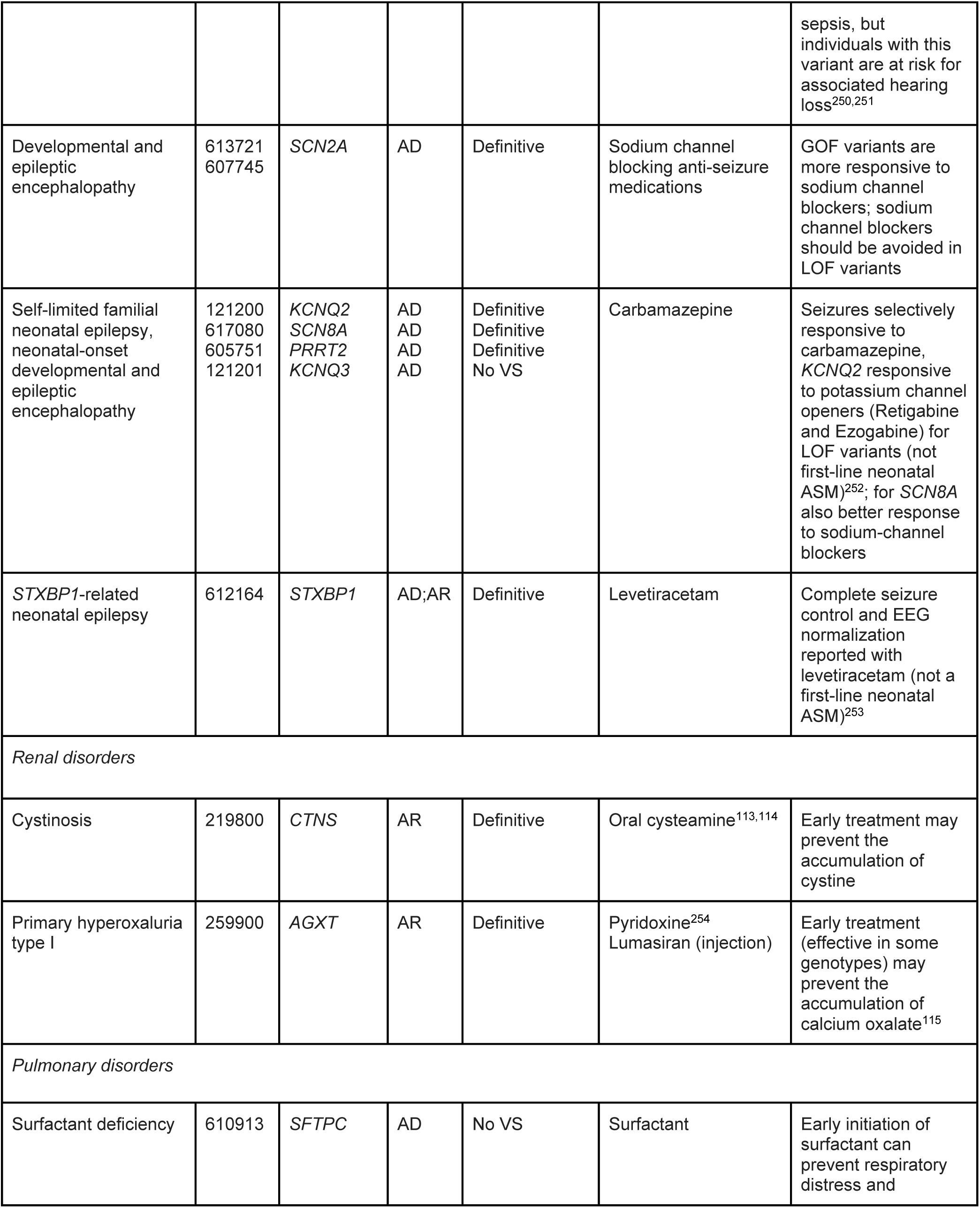
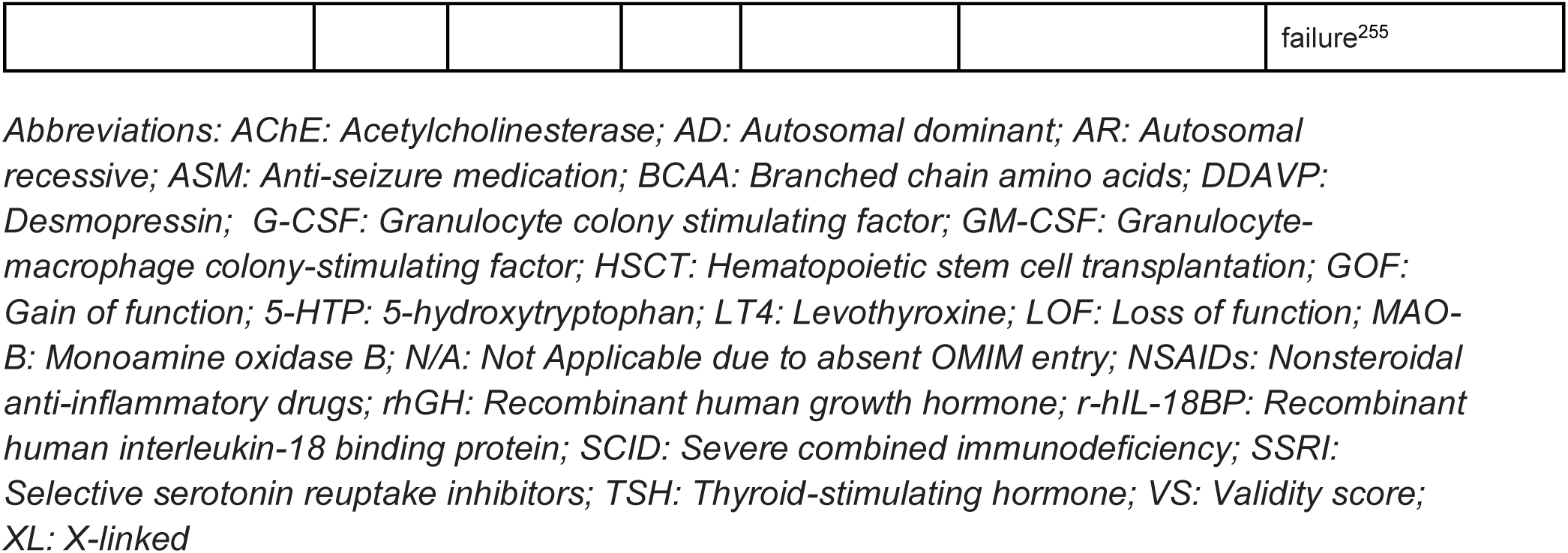
Genes associated with disorders with clinically available therapies that could be applied in the first week of life (n = 267).

### Inclusion criteria

#### Safety and efficacy of treatment

With any fetal therapy, two patients are implicated: the pregnant individual and the fetus.^35^ Because of this, the risk-benefit ratio of a fetal intervention must be tolerable, and there must also be potential efficacy without inflicting undue risk to either the pregnant person or fetus. For example, certain procedural fetal interventions carry a risk of preterm delivery, premature rupture of membranes, and oligohydramnios,^36^ while the risks of medication administration to the pregnant patient may vary.^37^ We defined a safe *in utero* therapy as one which did not result in fetal demise, did not lead to adverse side effects in the neonate or child, and did not cause unexpected adverse events in the mother.

Although some disease-targeted interventions have been successful, others have led to adverse events, and therefore were not included. For instance, intrauterine dexamethasone treatment for congenital adrenal hyperplasia (clinicaltrials.gov NCT02795871, NCT00617292) has extensive literature dating back to the 1980s and 1990s, but more recently was discovered to lead to cognitive impairment in children and was therefore deemed to be potentially harmful.^38–44^ Additionally, reports of significant maternal side effects from the dexamethasone administration present another concern.^45^

Disease efficacy was defined as improved neonatal outcomes when compared to the natural history of the disease.^46, 47–49^ We selected only disorders with treatments that are considered to be safe and potentially effective in human case series or clinical trials. For instance, the benefit of an intervention may be incomplete or inconsistent, such as a case report of maternal biotin administration to a fetus with holocarboxylase synthetase deficiency, in which the authors concluded that although it may have improved fetal growth, the prenatally administered dose was insufficient to prevent the neonatal acidotic crisis this particular patient experienced.^50^

For disorders with treatments that may improve clinical outcomes in the first week of life, the available treatments are broadly considered safe for affected infants. In many cases, a confirmatory non-molecular test, such as a biochemical laboratory test or flow cytometry, can be completed shortly after birth to determine if signs of disease are present. Such tests ensure that the appropriate treatment is applied only to infants who are symptomatic or have orthogonal evidence of disease, which is of particular importance for disorders with incomplete penetrance or variable expressivity.

#### Clinical severity

We selected only disorders that were associated with critical or chronic childhood illness. Additionally, the majority of the disorders included do not lead to structural anomalies. These disorders would therefore not be detected by sonography, nor would they likely be considered causative of a sonographic abnormality for which GS is ordered.

### Exclusion criteria

#### Gene-disease validity

Gene-disease validity refers to the strength of evidence supporting or refuting a claim that variation in a particular gene causes a corresponding monogenic disorder.^51^ All tables predominantly include genes with definitive, strong, or moderate gene-disease validity as annotated in ClinGen.^51^ Some genes have not yet been curated by ClinGen. Genes with limited, disputed, or refuted ClinGen validity scores have been excluded.

### Search strategy for treatable genetic disorders

A literature review was conducted to develop the lists of disorders that can be treated *in utero*. The authors conducted a review of clinicaltrials.gov (search terms: “fetal therapy”, “genetic” returned 256 results, with 8 relevant studies), ChatGPT^52^ and Microsoft CoPilot^53^ queries for available prenatal therapies for genetic disease, and a medical librarian-led search in Embase (search terms: “fetal therapy,” “fetal treatment,” “*in utero* therapy,” “*in utero* treatment,” “prenatal therapy,” “prenatal treatment” and limited to randomized controlled trials, clinical trials, case studies, and case reports) that yielded 593 titles and abstracts (Table S2) which were reviewed by two authors (J.L.C. and M.D.) for relevance. The searches were limited to English. These titles and abstracts were reviewed using Covidence.^54^ Articles deemed relevant by one or both authors were added to the tables and manuscript text if they met inclusion criteria. Full text manuscripts were reviewed on an as-needed basis.

To develop a list of disorders that are treatable in the first week of life, the authors reviewed the disorders on the Recommended Uniform Screening Panel (RUSP)^55^ and a list of 651 genes associated with treatable genetic disorders,^56^ aggregated from various online tools and published reports.^22,57–61^ Using these previously compiled lists, the authors included disorders that could benefit from antenatal detection and early postnatal treatment to improve neonatal outcomes. Additionally, the authors incorporated several disorders based on new evidence since the publication of the prior lists or as deemed appropriate according to the selection criteria.

Authors with expertise in various disease areas constructed each clinical section of this list (cardiac disorders: A.R.; endocrine disorders: D.M.M.; gastrointestinal disorders: A.S., J.R.T.; hematologic disorders: M.D.F.; inborn errors of immunity: R.H.; inherited metabolic disorders: R.G., N.B.G.; neurologic disorders: M.A.W.; renal disorders: W.T.). The list of all genes and disorders were then reviewed by the senior author (N.B.G.) for consistency with the selection criteria.

### Genetic disorders with *in utero* fetal therapies

#### Evidence for fetal diagnosis and fetal therapy

An increasing number of genetic disorders can now be treated in the fetal period through a range of therapeutic methods (Figure 1). The evidence for fetal treatment in humans includes case reports and clinical trial data.^62–65^

**Figure 1.**
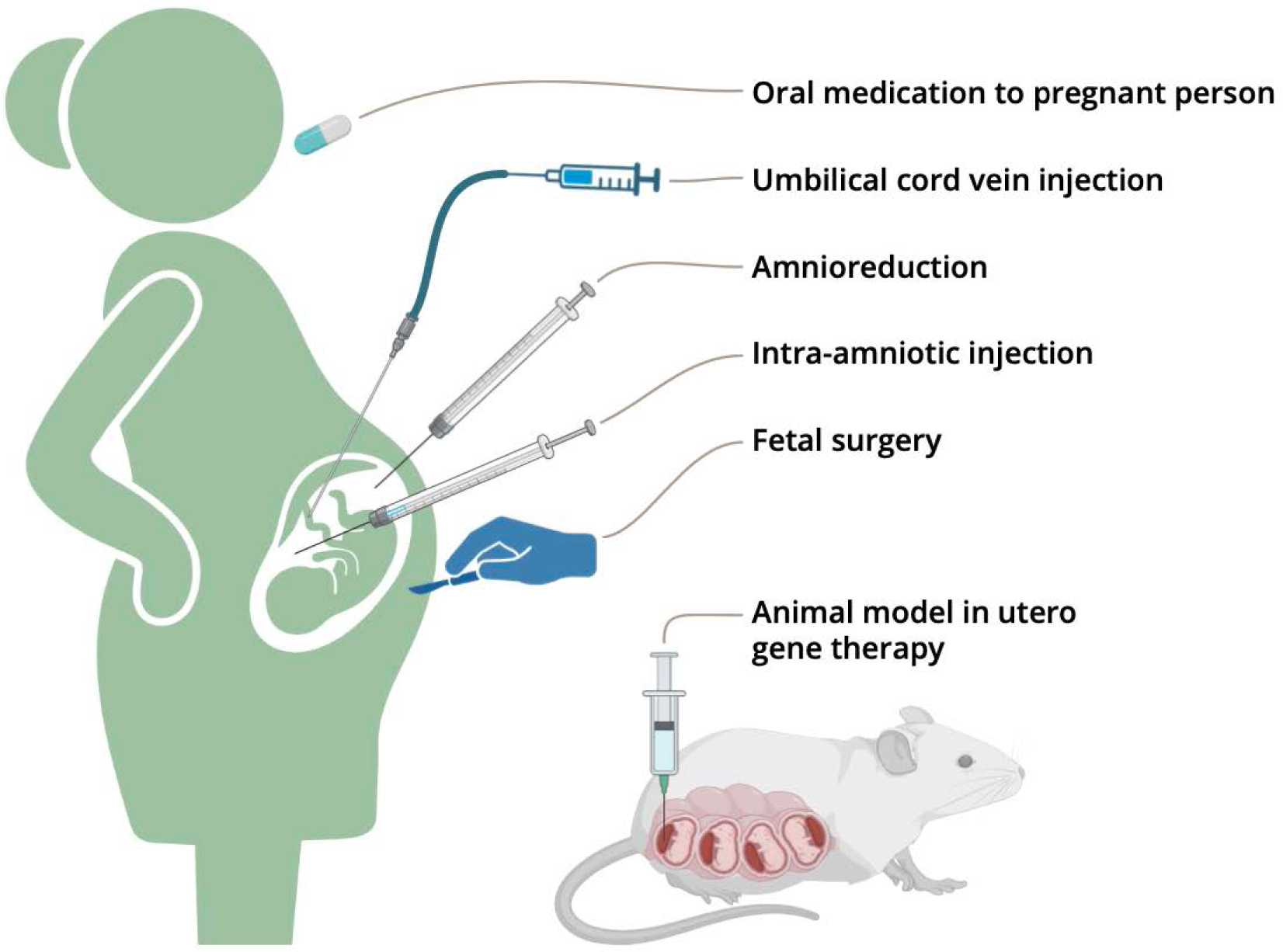
Methods of delivering fetal therapy in humans and animal models (created with Biorender).

##### Clinical trials

Systemically administered enzyme replacement therapy delivered through the umbilical cord vein has advanced to human clinical trials. This route of administration, also used for transfusions in fetal anemia,^66,67^ delivers therapy directly to the fetus and has an acceptable safety profile. Another human clinical trial involves intra-amniotic injection of the protein that is absent in X-linked hypohidrotic ectodermal dysplasia.^68^

Additionally, administering low-toxicity medications to a pregnant patient, either orally or through injections or infusions, can treat fetal genetic disorders by crossing the placenta. For instance, certain IMD can be treated with a nutritional supplement or medication provided to the mother, which then crosses the placenta, and improves enzyme activity or prevents toxic substrate accumulation in the fetus.^69–72^ Furthermore, promising outcomes have also been observed in more prevalent diseases such as cystic fibrosis, in which modulator therapy administered to pregnant patients can improve outcomes of affected fetuses.^47–49^ The ongoing exploration of therapeutic delivery to the pregnant patient or direct delivery to the fetus remains a viable path forward.

##### Case reports

Case reports for disorders such as *TRMU* deficiency and cobalamin C deficiency, in which a safe postnatal medication is trialed prenatally via administration to the pregnant person, provide early evidence from which larger prospective clinical trials can be launched.^69–72^ For these cases, the risk-benefit profile is favorable due to the low potential for maternal medication side effects. Among the case reports of oral administration of medication to a mother carrying an affected fetus, it is evident that the underlying molecular cause of a symptomatic presentation may help guide specific intervention. In a recent example, a case of fetal bradycardia with positive maternal autoimmune antibodies was found to be unresponsive to maternal dexamethasone treatment, and genetic testing later revealed a *KCNH2* variant-induced long QT syndrome, demonstrating how molecular diagnosis can guide management by alerting the team that conventional prenatal treatment for a common problem may not be sufficient.^73^ Relatedly, pathogenic variants in *SCN5A*, which can cause long QT syndrome type 3, can be treated with targeted medications.^73,74^ Additionally, there are times that a genetic diagnosis can lead to management changes such as avoiding the otherwise accepted prenatal therapy; for example, fetal chylothorax in fetuses with *PTPN11*-related Noonan syndrome responded poorly to *in utero* pleurodesis by OK-432.^75^

Disorders that require disease-specific delivery management should be considered for inclusion in an actionable fetal findings list as well. For instance, for fetuses affected by a hemophilia or a factor deficiency, cesarean section could be considered and use of vacuum-assisted delivery or forceps should be avoided to prevent intracranial hemorrhage.^76^

##### Preclinical research

Ongoing preclinical research in the field of fetal gene therapy explores various delivery routes and vectors across mouse, rat, and canine models.^62–65^ These delivery routes include injections that are intrahepatic, intracerebroventricular, intraplacental, intraperitoneal, intravenous, and into the yolk sac.^62^ While the field of *in utero* gene therapy and gene editing continues to hold great promise for a range of disorders, some approaches have been met with limited success, such as hematopoietic stem cell gene therapy in a canine model of MPS I, which was unable to reduce disease burden.^77^

### Disorders for which prenatal genomic diagnosis may improve outcomes in the first week of life

Although NBS has prevented morbidity and mortality in infants with a range of IMD and other genetic disorders, several disorders included on the RUSP can cause critical illness in the first week of life before NBS results are typically returned. Additionally, there are a range of other disorders that are not yet included in public health NBS programs which also present with symptoms or have clinical therapies that could be initiated promptly after birth.

#### Evidence for treatment in the first week of life

##### Cardiac disorders

The genes that most commonly account for long QT syndrome can cause fatal arrhythmias in fetuses or infants, and have even been implicated in some cases of infant and childhood sudden death.^78,79^ Medical management with beta blockers or the implantation of a cardioverter-defibrillator may be lifesaving. Additional genes associated with long QT syndrome account for a very small proportion (<1%) of diagnoses.^80^

##### Endocrine disorders

Many neonatal-onset endocrine disorders can cause electrolyte disturbances, hypoglycemia, or salt-wasting crises in the first days of life, which lead to neurologic sequelae and even death.^81,82^ While these symptoms can be partially managed without knowledge of the specific genotype, prenatal detection of these disorders may allow for more proactive clinical care and thereby prevent acute manifestations.^83^ Additionally, in the example of congenital hyperinsulinism, knowledge of the underlying genotype allows for the appropriate treatment to be expedited, as diazoxide may not be effective in certain genetic subtypes.^84^ Relatedly, the more common forms of neonatal diabetes (*KCNJ11*, *ABCC8*) typically present after the first week of life, but can be improved by targeted treatment with sulfonylureas.^85^

##### Gastrointestinal disorders

There are several gastrointestinal disorders for which early identification could improve outcomes in infants. A variety of congenital diarrheas and enteropathies manifest immediately after birth and can be treated with disease-specific fluid and electrolyte therapies in the first few days of life.^86,87^ Importantly, limitation of enteral feeding can lead to life-threatening acid-base instability.^88^ Additionally, accurate identification of these disorders may prevent potentially unnecessary evaluations or surgeries for conditions such as pseudo-obstruction, which show similar imaging findings to certain genetic conditions.^89^ Autosomal recessive hyperlipoproteinemia, type 1D, can present with chylomicronemia shortly after birth, and early intervention can reduce morbidity.^90^

##### Hematologic disorders

A wide variety of genetic disorders of hematopoiesis as well as plasma proteins, particularly those involved in hemostasis, are amenable to *in utero* or perinatal therapies or management strategies. Nearly all severe fetal anemias, for example, respond to *in utero* transfusion, which can bridge the gap to birth, after which chronic transfusion, hematopoietic stem cell transplantation, or, increasingly, gene therapy can be delivered.^91–98^ The erythroid porphyrias, erythropoietic protoporphyria and congenital erythropoietic porphyria, do not themselves cause severe anemia, but the overproduction of porphyrins results in extreme light sensitivity, warranting the avoidance of neonatal phototherapy.^99^ Disorders of granulocyte or platelet numbers or function generally do not cause disease in pre- or perinatal life, but early identification may lead to prophylactic therapies to avoid bleeding or infectious complications.^100–102^ Recognition of clotting factor deficiencies may be an indication for cesarean section, prompt initiation of factor replacement, or avoidance of common procedures such as circumcision.^76,103^

##### Inborn errors of immunity

NBS for severe combined immunodeficiencies (SCID) has improved morbidity and mortality for infants with the most severe form of immunodeficiency. Nonetheless, neonates with SCID can acquire life-threatening infections, in particular cytomegalovirus (CMV) from breast milk, prior to detection by NBS and confirmatory flow cytometry testing.^104,105^ In addition, false negative NBS results for SCID do occasionally occur.^104^ Prenatal detection of fetuses at risk for SCID would allow for improved management both pre and post-natally. Prenatally, families could be referred for initial bone marrow transplant evaluation and HLA typing could be initiated. Families could be referred to specific tertiary centers with providers experienced with treating patients with SCID. In the neonatal period, measures including isolation precautions, immune prophylaxis, and counseling against breastfeeding for mothers positive for CMV could further reduce morbidity and mortality as confirmatory testing is performed. Of note, we did not include genes recently associated with SCID which have limited or not yet curated gene-disease relationship based on the ClinGen SCID-CID expert panel, e.g. *MAN2B2* and *BCL11B*, both of which are associated with congenital anomalies and could be ascertained by indication-based testing.^106^

Variable expressivity is a common feature of inborn errors of immunity (IEIs), including SCID and combined immunodeficiencies (CIDs). Many genes are associated with both SCID and CIDs, often due to hypomorphic variants linked to the latter. Due to the critical importance of identifying patients at risk for SCID, we included genes associated with CIDs that can present in the neonatal period with SCID.^107^ Because of the heterogeneity of CIDs and marked variable expressivity, it is challenging to definitively distinguish which CIDs would benefit from diagnosis within the first week of life. Early detection of many IEIs including CIDs could decrease morbidity and mortality during infancy, but is beyond the scope of this review.^56^

We included genes associated with agammaglobulinemia, a category of disorders predominantly associated with antibody deficiencies. Although infants with agammaglobulinemia typically present at 3-6 months of age, earlier diagnosis and treatment would likely prevent morbidity and mortality.^105^ Several countries are implementing B cell kappa chain receptor excision circles (KRECs) screening in conjunction with SCID NBS. This screening can detect patients at risk for agammaglobulinemia.^105^ These programs will provide critical information regarding the utility of neonatal detection which can help inform genes chosen for the actionable fetal findings list.

We also included genes associated with diseases of immune dysregulation, congenital defects of phagocyte number, function, or both, defects in intrinsic and innate immunity and autoinflammatory disorders. We included IEIs that can present during the neonatal period and are treatable.^107,108^ We did not include complement deficiencies, as the typical age of onset is usually in childhood, although one neonatal presentation has been reported.^107,108^ This is an area of active investigation, in particular for diseases of immune dysregulation and autoinflammatory disorders.^108^ We anticipate that the list of genes associated with fetal and perinatal presentation will rapidly increase as these disorders are further characterized with growing awareness of potential fetal presentation.

Variable expressivity and incomplete penetrance are common features of IEIs. In particular, for IEIs with genotype-phenotype associations, consideration of both gene and specific variant would be important to consider prior to inclusion in an actionable fetal findings list. Neonatal orthogonal testing for IEIs, in particular flow cytometry, is a powerful additional tool when penetrance is unknown.

##### Inherited metabolic disorders

Many IMD lead to symptoms in the first days of life, prior to the receipt of NBS results. The detection of at-risk fetuses would allow pregnant patients to prepare for delivery in a clinical center where a specialized biochemical genetics team is present, or allow advanced notice for the birth center to procure the necessary metabolic medications and formulas.

In a recent example, a female fetus with pyruvate dehydrogenase deficiency was diagnosed via exome sequencing in the setting of structural brain malformations.^109^ This prenatal diagnosis allowed for interdisciplinary delivery planning among the pregnant patient’s obstetric providers and the institution’s pediatrics teams, including biochemical genetics specialists, neonatologists, and dieticians with expertise in the ketogenic diet. The infant was placed on a ketogenic diet immediately after birth, and as a possible result, experienced no seizures or lactic acidosis in the neonatal period. This case illustrates the potential opportunity for infants with other IMD requiring specialized diets or medications, such as organic acidemias or urea cycle disorders, to receive appropriate treatment beginning at birth. In some cases, prompt initiation of treatment may prevent the accumulation of toxic intermediates that lead to severe metabolic decompensations, characterized by lethargy, seizures, and even early death. Early treatment of IMD therefore has the potential to improve lifelong health and quality of life for affected individuals.

Of note, one challenge that may arise across these disorders is that specific variants in some genes (such as those associated with the myopathic form of carnitine palmitoyltransferase type II deficiency) are strongly associated with attenuated or late-onset forms of disease, which do not meet the inclusion criteria for this review. If these genes were adapted for an actionable fetal findings list, further discussion will be needed regarding which PLPV to analyze and report.

##### Neurologic disorders

The management of infants at risk for many monogenic disorders with neurologic symptoms could be improved by identification prior to birth. In particular, several syndromes causing neonatal seizures are optimally treated with specialized management that differs from the standard of care; prompt initiation of the appropriate anti-seizure medication is more likely to lead to complete seizure control.^110^ Additionally, for disorders such as the congenital myasthenia syndromes, the effect of acetylcholinesterase therapy depends directly on the genotype, which in some cases can worsen symptoms and lead to critical illness.^111^

One unique entity that meets criteria for inclusion is the mtDNA variant m.1555A>G, a risk allele for aminoglycoside-induced deafness. Although this variant is not expected to cause symptoms *a priori*, for infants who undergo preventive treatment of sepsis, commonly-used aminoglycoside antibiotics may put them at risk for hearing loss. Instead, targeted pharmacologic treatment for these infants might lead to the use of a different antibiotic.^112^

##### Renal disorders

Infants at risk for two renal disorders, cystinosis and primary oxaluria, would particularly benefit from treatment in the first week of life to preserve renal function. Cystinosis leads to cystine accumulation in various tissues, including the kidney. Although clinical symptoms do not typically occur until approximately 6 months of age, glomerular damage accumulates from birth. Early treatment with cystine-depleting agents such as cysteamine immediately after birth can slow this damage.^113,114^ Primary oxaluria, which presents in the first months of life in 10% of affected individuals, results in oxalate accumulation, causing nephrocalcinosis, nephrolithiasis, and progressive kidney damage.^115^

## Discussion

Fetal GS is poised to play an increasingly significant role in prenatal diagnosis. The ISPD recommends offering GS to all pregnant patients whose fetuses have structural abnormalities,^1^ which would encompass up to 2-3% of pregnancies.^2^ Currently, no established guidelines regarding which monogenic variants to report in fetal GS exist and the role of secondary findings remains unclear. Importantly, parents have demonstrated interest in using GS to diagnose disorders with available experimental *in utero* therapies. In surveys of parents with children affected by mucopolysaccharidoses, for example, the majority of parents had a favorable attitude toward phase 1 clinical trials for fetal therapy.^116^ Similarly, survey results from families affected by spinal muscular atrophy and sickle cell disease overwhelmingly supported prenatal diagnosis, and the majority expressed an interest in fetal therapy.^117,118^ Identifying treatable monogenic disorders during the fetal period could therefore enhance patients’ care options and autonomy during pregnancy, as well as improve the neonatal and lifelong health of affected infants.

In this integrative review, we compiled a list of 295 genes associated with disorders for which therapeutic intervention—either in the fetus (53 genes) or in the first week of life (267 genes, including 25 genes that appear on both lists)—could improve health outcomes. At present, we suggest that all disorders for which there have been successful treatments reported in individual human cases, disorders with fetal treatments that are currently in clinical trials, and all disorders for which treatment in the first week of life may improve outcomes should be considered for an actionable fetal findings list. This list could be offered to pregnant patients undergoing diagnostic fetal GS, or as exome sequencing becomes feasible on cfDNA, as a standalone non-invasive screening tool.^30,119^ Of note, however, the list in this review is not comprehensive nor consensus-based and will require updates as new research and therapies emerge.

Although GS allows for querying the entire genome, and any disease-associated genomic finding could be considered actionable in the perinatal period, presenting the option to assess a list of actionable fetal disorders is an important step toward enhancing the autonomy of pregnant patients. In the future, pregnant patients could be offered the option for GS to focus on variants associated with sonographic findings, or to also include additional actionable findings, or analysis of the whole genome. If whole genome analysis were pursued, these lists of disorders could also be used to guide discussions on potential treatment opportunities, which would provide patients with a more comprehensive range of care options. Furthermore, reporting all PLPV associated with childhood-onset disorders may be premature and pose challenges to patient counseling, as the penetrance and expressivity of many variants are not yet well-understood.^120^

Although the treatable disorders listed in this review are all relatively well-characterized in the medical literature, other challenges remain in establishing the clinical utility of an actionable fetal findings list. The penetrance of many of these disorders is unknown and the paucity of diagnostic imaging signs or non-molecular confirmatory testing available in fetuses limits diagnostic certainty.^121,122^ Fetal GS is complicated by incomplete phenotyping due to ultrasound limitations^14^ and many disorders may have no discernable prenatal phenotype to substantiate the diagnosis.^121,122^ Efforts are underway to expand entries in the Human Phenotype Ontology related to prenatal presentations; however, this does not address incomplete phenotyping related to technological limitations.^122^ While tests such as fetal enzyme activity using placental^123^ or umbilical cord blood samples are possible, interpreting results is difficult without established fetal norms.^124^ After birth, genetic diagnoses can be confirmed by non-genetic findings, but during the fetal period, treatment decisions may rely solely on genetic information.

Implementation of a fetal actionable findings list may also be complicated by barriers to care in resource-limited settings and by psychosocial ramifications. Access to fetal GS is inequitable^125^ and the additional analysis of treatable disease genes could potentially widen health disparities. Reporting only PLPV, as has been recommended by the ACMG, may lead to inequities for reproductive couples of non-European ancestry in whom variants of uncertain significance are more common.^126^ When a positive finding is identified, it may be infeasible for some individuals to receive care at a clinical center where the appropriate treatment is available.^127^ Pregnant patients offered GS in the setting of a fetal anomaly might face an overwhelming amount of information if offered multiple secondary or actionable findings lists.^128–131^ As the clinical use of fetal GS expands, the ethical, legal, and social ramifications of this technology will continue to be a field of active research.

The use of trio GS as a tool to investigate conditions affecting the pregnant patient is also an important new direction for investigation. Given that fetal GS typically includes a sample from the pregnant patient and that the maternal genome is also assessed in cfDNA sequencing, additional genetic disorders that affect fetal or maternal health, or lead to pregnancy complications, could also be considered for inclusion. For example, the pregnant patient’s sample could reveal disorders that are teratogenic to the fetus, such as maternal phenylketonuria^132^ or thrombotic thrombocytopenic purpura.^133^ Additionally, analysis of this sample could identify a risk for disorders that may present in the breastfeeding infant, such as transient neonatal zinc deficiency or acrodermatitis enteropathica,^134^ which can be prevented by supplementation of the deficient nutrient. Furthermore, identifying genetic conditions that affect the health of the pregnant or the postpartum patient, such as ornithine transcarbamylase deficiency, vascular Ehlers-Danlos syndrome, or cardiomyopathies, could prevent deadly complications such as hyperammonemia or uterine rupture.^135^ Lastly, detecting fetal conditions like fatty acid oxidation disorders, may also inform prenatal care, as they can cause secondary effects in the pregnant patient.^136^

As access to fetal GS grows and the capabilities of cfDNA sequencing advance, the field of prenatal genetic diagnosis will continue to expand. The implementation of an actionable fetal findings list has the potential to enhance the autonomy of pregnant patients and improve the health of infants with rare diseases. There is evidence to suggest that a large number of genes are associated with conditions that are treatable *in utero* or the immediate perinatal period. In time, our understanding of variant curation and prenatal phenotypes will grow, which will improve post-test counseling for pregnant patients with PLPVs found on fetal GS. Although challenges remain regarding the equitable implementation of fetal GS, an actionable fetal findings list could currently be offered to individuals who are undergoing this test, and eventually may form the basis of a non-invasive screening tool performed on cfDNA that could be offered to all pregnant patients.

## Data Availability

All data produced in the present work are contained in the manuscript.

## Acknowledgements

The authors would like to thank Leila Ledbetter for conducting the Embase search on prenatal therapies and Eric Monson for his assistance with Figure 1.

## Data and code availability

This study did not generate any datasets or code.

## Declaration of generative AI and AI-assisted technologies in the writing process

During the preparation of this work the author(s) used Open AI ChatGPT and Microsoft CoPilot in order to generate ideas about disorders with fetal therapies and clarify grammar and syntax of some sentences. After using this tool/service, the author(s) reviewed and edited the content as needed and take(s) full responsibility for the content of the publication.

