## Supplemental Tables for "Advancing precision care in pregnancy through an actionable fetal findings list"

### Table S1. Genes associated with disorders with clinically available and experimental *in utero* fetal therapies in animal models (n = 17).

| | | | | | | | |
|---|---|---|---|---|---|---|---|
| *Animal models* | | | | | | | |
| Hypophosphatasia | 241500 | *ALPL* | AR | Definitive | AAV gene therapy [1–3] | Animal model | Endocrine disorder |
| Beta Thalassemia | 613985 | *HBB* | AR | Definitive | *In utero* stem cell transplantation [4] | Animal model | Hematologic disorder |
| Hemophilia A | 306700 | *F8* | XL | Definitive | Administration of human amniotic fluid mesenchymal stromal cells (hAFMSCs) via *in utero* transplantation [5] | Animal model | Hematologic disorder |
| Sickle cell disease | 603903 | *HBB* | AR | Definitive | *In utero* stem cell transplantation [4,6] | Animal model | Hematologic disorder |
| Citrullinemia type 1 | 215700 | *ASS1* | AR | Definitive | Gene therapy[7] | Animal model | Inherited metabolic disorder |
| Crigler–Najjar type 1 | 218800 | *UGT1A1* | AR | No VS | Gene therapy[8,9] | Animal model | Inherited metabolic disorder |
| Hereditary tyrosinemia type 1 | 276700 | *FAH* | AR | Definitive | Gene therapy[10] | Animal model | Inherited metabolic disorder |
| Human erythrocyte R-type pyruvate kinase deficiency | 266200 | *PKLR* | AR | Definitive | Gene therapy[11] | Animal model | Inherited metabolic disorder |
| Tay-Sachs disease | 272800 | *HEXA* | AR | Definitive | Gene therapy[12] | Animal model | Inherited metabolic disorder |
| *RPE65*-related disease | 613794<br>618697<br>204100 | *RPE65*<br>*RPE65*<br>*RPE65* | AR<br>AD<br>AR | Definitive<br>Strong<br>No VS | Subretinal injection of | Animal model | Inherited retinal disorder |

| | | | | | | | |
|---|---|---|---|---|---|---|---|
| | | | | | AAV gene therapy[13] | | |
| RASopathies, specifically Noonan syndrome | 609942 | *KRAS* | AD | Definitive | Exposure of pregnant K-*Ras* V14I mice to a MEK inhibitor[14] | Animal model | Multi-system disorder |
| Angelman syndrome | 105830 | *UBE3A* | AD | Definitive | Anti-sense oligonucleotide (ASO)[15] | Animal model | Neurologic disorder |
| Duchenne Muscular Dystrophy | 310200 | *DMD* | XL | Definitive | Gene editing of genomic DNA using single-stranded oligodeoxy-Nucleotides in MDX animal model[16] | Animal model | Neurologic disorder |
| Genetic hearing loss | 613718<br>602092 | *MSRB3*<br>*USH1C* | AR<br>AR | Definitive<br>Definitive | *In utero* gene therapy[17,18]; ASO into amniotic cavity for USH1C[19]; *in utero* gene therapy for MSRB3[20] | Animal model | Neurologic disorder |
| Neurodevelop-mental conditions (i.e. Rett syndrome) | 312750 | *MECP2* | XL | Definitive | Theoretical use of SAMe, choline, and valproic acid as possible epigenetic drugs[21] | Animal model[22] | Neurologic disorder |
| Spinal muscular atrophy | 253300 | *SMN1* | AR | No VS | AAV gene therapy[23]; maternal risdiplam (to pregnant dams)[24] | Animal model | Neurologic disorder |
| Surfactant deficiency | 610913 | *SFTPC* | AD | No VS | *In utero* gene editing[25,26] | Animal model | Pulmonary disorder |

*Abbreviations: AAV: adeno-associated virus; AD: autosomal dominant; ASO: antisense oligonucleotide; AR: autosomal recessive; ERT: enzyme replacement therapy; HSC: hematopoietic stem cells; LXR-agonist: Liver X receptor agonist; SAMe: S-adenosylmethionine; VS: validity score; XL: X-linked*

**Table S2. Search strategies and results for titles and abstracts detailing genetic disorders with *in utero* therapies.** Date: August 13, 2024. Database / Study Registry (including vendor/platform): Embase (Elsevier).

| Set # | Search Strategy | Results |
|---|---|---|
| 1<br><br>*Fetal therapy* | 'fetal therapy'/exp OR 'fetal therapy' OR 'fetal therapies' OR 'foetal therapy' OR 'foetal therapies' OR 'fetal treatment' OR 'fetal treatments' OR 'in utero treatment' OR 'in utero treatments' OR 'in utero therapy' OR 'in utero therapies' OR 'prenatal therapy' OR 'prenatal therapies' OR 'prenatal treatment' OR 'prenatal treatments' | 5,610 |
| 2<br><br>*And study filters* | ('fetal therapy'/exp OR 'fetal therapy' OR 'fetal therapies' OR 'foetal therapy' OR 'foetal therapies' OR 'fetal treatment' OR 'fetal treatments' OR 'in utero treatment' OR 'in utero treatments' OR 'in utero therapy' OR 'in utero therapies' OR 'prenatal therapy' OR 'prenatal therapies' OR 'prenatal treatment' OR 'prenatal treatments') AND ( | 1,041 |
| 3<br><br>*And article status and english* | ('fetal therapy'/exp OR 'fetal therapy' OR 'fetal therapies' OR 'foetal therapy' OR 'foetal therapies' OR 'fetal treatment' OR 'fetal treatments' OR 'in utero treatment' OR 'in utero treatments' OR 'in utero therapy' OR 'in utero therapies' OR 'prenatal therapy' OR 'prenatal therapies' OR 'prenatal treatment' OR 'prenatal treatments') AND ([controlled clinical trial]/lim OR [randomized controlled trial]/lim OR 'clinical trial' OR 'controlled trial' OR 'case report' OR 'case study') AND ([article]/lim OR [article in press]/lim OR [data papers]/lim OR [letter]/lim OR [note]/lim OR [review]/lim OR [short survey]/lim OR [preprint]/lim) AND [english]/lim | 601 |
